# Symptom variability and treatment response in pernicious anaemia: a longitudinal daily symptom tracking study

**DOI:** 10.64898/2026.11.03.26367157

**Authors:** Alfie Thain, Kathryn Hart, Katrina Burchell, Petra Visser, Kourosh R Ahmadi

**Affiliations:** School of Biosciences and Medicine, University of Surrey, Guildford, UK; The Pernicious Anaemia Society, Ipswich, UK

## Abstract

Pernicious Anaemia is an autoimmune-mediated disorder of vitamin B_12_ absorption, managed using fixed-interval vitamin B_12_ replacement regimens. Despite treatment, many patients report persistent or recurrent symptoms, suggesting substantial inter-individual variability in treatment response. More tailored approaches may therefore be needed to reduce symptom burden and improve quality of life. This prospective observational study used high-frequency longitudinal digital symptom monitoring to characterise symptom dynamics and heterogeneity in treatment response.

UK adults with pernicious anaemia, recruited via the Pernicious Anaemia Society, self-reported daily symptoms (severity 0-10) and vitamin B_12_ treatments over three months. Analyses quantified within- and between-person variability, injection-aligned symptom response kinetics, and differences in treatment response by injection frequency.

Sixty-eight participants were included in the longitudinal analyses, providing >5,100 person-days of symptom data over a mean of 76 days (SD 28). Most symptom variance was attributable to stable between-person differences (ICC range, 0.58-0.80), indicating the persistence of individual symptom phenotypes. Vitamin B_12_ injections were followed by rapid symptom improvement, although magnitude and timing varied markedly by symptom domain. Seventy-one percent of participants were classified as responders for composite symptom burden (median 4.65 to 2.36; p<0.001), while 29% showed no meaningful multisystem improvement (3.59 to 3.58; p=0.63). Cognitive symptoms improved earliest, typically within 24-72 hours; fatigue and pain responses peaked at 3-5 days, and neuro-sensory symptoms at 4-6 days. Responses clustered within individuals, indicating distinct responder phenotypes. Overall symptom burden improved most among participants receiving monthly injections, whereas those injecting weekly or more often had higher pre-injection burden and smaller improvements.

These findings suggest that fixed-interval vitamin B_12_ replacement may not adequately reflect individual variation in symptom recurrence and treatment response. High-frequency digital symptom monitoring may provide a practical framework for characterising these patterns and for evaluating more individualised, symptom-guided vitamin B_12_ management.

**Author Summary:** Pernicious anaemia is a lifelong autoimmune condition that prevents the body from properly absorbing vitamin B_12_. Although regular vitamin B_12_ injections are used to treat the condition, many people continue to experience symptoms between injections and it is unclear why treatment works better for some people than others.

We asked 68 people with pernicious anaemia to record their symptoms and vitamin B12 treatments every day for up to three months. This allowed us to examine how symptoms changed over time and around individual injections. We found that symptoms often improved after vitamin B_12_ injections, but the size and timing of improvement varied considerably between people and between different symptoms. Some participants showed improvement across many symptoms, while others showed little change. We also found that symptoms often returned between injections, even in people receiving long-term treatment.

Our findings show how frequent digital symptom tracking can reveal patterns that may be missed during short clinical appointments. In the future, this approach could help clinicians and researchers evaluate whether vitamin B_12_ treatment can be better tailored to an individual’s symptoms and treatment response.

## Introduction

Pernicious Anaemia (PA) is a life-long, autoimmune disease characterised by the destruction of gastric parietal cells, leading to impaired vitamin B_12_ (B_12_) absorption [1], [2]. PA remains an overlooked condition associated with substantial diagnostic delay or misdiagnosis and *one-size-fits-all* management approaches [3], [4]. In the United Kingdom, maintenance treatment typically consists of fixed-interval intramuscular hydroxocobalamin injections every two or three months [3], [5]. However, more than half of patients seek more frequent treatment, either through their GP or often self-administered, to achieve better symptom control [4], [6].

People with PA report a wide range of neurological, cognitive, mood-related and systemic symptoms, including paraesthesia, balance problems, memory and concentration difficulties, fatigue, gastrointestinal symptoms, depression and anxiety [2], [7], [8], [9], [10], [11]. Many of these symptoms are non-specific and may also occur in other autoimmune, neurological or nutritional conditions, making attribution to pernicious anaemia challenging. Importantly, some individuals continue to experience persistent or recurrent symptoms despite long-term vitamin B_12_ replacement. The optimal approach to long-term maintenance treatment remains uncertain. Fixed-interval regimens are widely used, yet many patients report symptom recurrence before their next scheduled injection and seek more frequent treatment. It also remains unclear whether persistent symptoms reflect inadequate treatment, irreversible consequences of previous B12 deficiency, co-existing conditions, or other factors. The James Lind Alliance Priority Setting Partnership identified improving management and treatment as a key research priority in PA [5]. Addressing this priority requires a better understanding of what symptoms arise in PA, how each symptom varies between and within individuals and importantly if and to what degree these symptoms respond to B_12_ treatment.

### Rationale for Daily Symptom Tracking

Daily digital symptom tracking offers a promising approach to capturing within- and between-individual variability in PA symptomatology that is not detectable through conventional, cross-sectional designs. In rheumatoid arthritis, endometriosis, menstrual health and multiple sclerosis, digital symptom monitoring has characterised heterogeneous symptom trajectories, informed clinical decision-making and supported more effective personalised management strategies [12], [13].

Symptoms in PA are central to daily functioning but can vary markedly in their severity, timing and responsiveness to B12 therapy between individuals. Existing studies have relied largely on cross-sectional or retrospective assessments, limiting understanding of short-term symptom fluctuations and treatment-aligned response dynamics [3], [7], [10], [14]. Longitudinal daily monitoring across multiple treatment cycles provides an opportunity to characterise within-person symptom trajectories, identify treatment-response phenotypes, and evaluate whether fixed-interval maintenance regimens adequately reflect the heterogeneity of symptom response observed in clinical practice. This is consistent with current NICE guidance, which recommends adjusting maintenance treatment according to symptom severity [15].

In the current study, we aimed to characterise symptom dynamics and treatment response among individuals living with PA using longitudinal daily digital symptom tracking. Specifically, we sought to: (1) characterise symptomatology presentation and burden across multiple B_12_ treatment cycles; (2) quantify within- and between-individual variability in symptom severity; (3) identify treatment-response phenotypes; and (4) evaluate the impact of standard versus non-standard treatment frequencies on symptom trajectories. We identified substantial between-person variation in symptom burden, alongside distinct injection-aligned patterns and individual differences in treatment response.

## Materials and Methods

### Survey design and setting

This study used an observational, prospective daily symptom-tracking design delivered as an online questionnaire hosted on Qualtrics. The survey was developed by the University of Surrey research team in collaboration with the PAS to ensure relevance and feasibility for daily completion. Participants were recruited between April 2023 and May 2023. Data collection took place between June 2023 and October 2023. Participants were invited to complete the survey once per day for up to three months, providing repeated measures of B_12_ treatment, symptom presentation and severity over time.

All participants provided electronic written informed consent prior to participation via an online consent form integrated within the Qualtrics platform. The study was conducted remotely, with participants completing surveys on personal devices.

### Participants and recruitment

Participants were recruited via the PAS research repository, a database of individuals who had previously expressed interest in participating in research related to PA [4]. Eligible individuals were invited by email and provided with a participant information sheet, electronic consent form and a link to the baseline Qualtrics survey.

Automated daily reminder emails containing the survey link were sent throughout the participation period to support engagement and minimise missing data.

As this was an exploratory prospective observational study, no formal a priori sample-size calculation was performed. Recruitment was based on feasibility and the number of eligible participants available through the PAS research repository during the study period. Participants who withdrew from the study were excluded from the longitudinal analyses.

Prospective daily reporting and automated reminders were used to minimise recall bias and missing observations. Injection-level observations were aggregated within participants before cohort-level analyses to reduce disproportionate weighting of participants receiving more frequent injections.

### Eligibility criteria

Participants were eligible to take part if they met all of the following criteria:

- Were aged 18 years or older
- Self-reported a diagnosis of PA, and indicated that their diagnosis had been supported by at least one of the following: positive intrinsic factor autoantibodies, a positive Schilling test (historical), or endoscopic/histological evidence of autoimmune gastritis (AIG)
- Reported current treatment with intramuscular or subcutaneous B_12_ injections. Participants could also use additional B_12_ preparations, including oral tablets, sublingual formulations, sprays, or patches.
- Reported experiencing symptoms attributed to PA;
- Were resident in the United Kingdom;
- Had access to the internet and a personal device (mobile phone, tablet, or computer) to complete the online survey.

### Daily survey instrument and measures

Participants completed a brief daily online questionnaire capturing B_12_ treatment exposure, symptom presence and severity, and overall wellbeing (**Supplementary Table 1**).

Each day, participants reported whether they had received any B_12_ treatment and, if so, specified the route of administration (intramuscular injection, subcutaneous injection, oral, sublingual, spray, or patch) and formulation. Participants then reported symptom presence and rated severity from a list derived from prior work (**Supplementary Note 1**). Symptom severity was recorded using an 11-point numerical rating scale (0-10), where 0 indicated symptom absence and 10 indicated maximal severity. For each symptom, participants were provided with standardised quality-of-life and functional impairment anchored severity definitions:

• **Mild symptom**: The symptom is noticeable but doesn’t interfere with daily living activities, you get used to it and adapt to the symptom.

• **Moderate symptom**: The symptom is distracting throughout the day but can be ignored for some time when focusing on daily activities.

• **Severe symptom**: The symptom significantly interferes with daily activities and cannot be ignored at all.

Overall wellbeing was recorded on a 10-point scale from 1 (excellent) to 10 (poor), with lower scores indicating better wellbeing, and was analysed separately from symptom burden. Within-day items were compulsory, although participants could miss days and resume subsequently. A baseline questionnaire collected demographic and clinical characteristics, including age, sex, age at PA diagnosis, duration of symptoms before diagnosis, family history of PA, and co-existing autoimmune conditions.

The survey required approximately five minutes to complete. Participants were invited to complete a post-study feedback survey.

### Data management

Each participant was assigned a unique identifier to enable longitudinal linkage while preserving anonymity. Survey data were stored on the Qualtrics platform and exported to Microsoft Excel and R for cleaning and analysis following study completion. Missing daily entries were retained as missing and not imputed.

### Statistical analysis

All analyses were conducted in R (version 2023.12.0). Symptom severity scores were treated as ordinal repeated measures and analysed using non-parametric tests. Analyses were structured to: (i) describe baseline symptom burden and injection patterns, (ii) estimate within-person symptom change following B_12_ injections, (iii) quantify between-participant heterogeneity, and (iv) examine symptom clustering and domain-level response patterns.

Participants contributing <20% of expected daily entries were excluded from longitudinal analyses. Participants reporting no B_12_ treatment during follow-up were excluded from treatment-effect analyses. Injection-cycle analyses included participants contributing ≥1 eligible isolated injection with paired pre- and post-treatment windows.

Within-person treatment effects were estimated using temporally aligned three-day pre-injection (t-3 to t-1) and three-day post-injection (t+1 to t+3) windows, restricted to injections isolated from adjacent doses. To avoid over-representation of frequent injectors, injection-level summaries were aggregated to participant-level mean pre- and post-treatment severities prior to cohort-level analyses. Paired changes were summarised using medians and interquartile ranges and tested using Wilcoxon signed-rank tests.

For injection-frequency analyses, participants were grouped according to their predominant injection schedule: 2-3 monthly, monthly, every 2-3 weeks, or weekly or more often. For comparisons of standard versus non-standard treatment frequency, 2-3 monthly treatment was classified as standard and all more frequent schedules as non-standard.

Intraclass correlation coefficients (ICCs) were calculated to partition within- and between-participant variance. Symptom co-occurrence was assessed using Spearman’s rank correlation matrices. Treatment responder status was classified using pre-specified absolute and percentage change thresholds informed by IMMPACT and minimal clinically important difference (MCID) criteria [16], [17], [18], [19], [20]. Substantial improvement was defined as ≥50% or ≥3-point reduction; clinically important improvement as 30-49% or 2-2.99-point reduction; minimal improvement as 10-29% or 1-1.99-point reduction; non-response as <1-point and ±10% change; and worsening as ≥10% increase. Person-level clustering of response was examined using multilevel logistic regression models with random intercepts for participants and symptoms.

Exploratory analyses examined associations between predominant responder status and age, sex, age at PA diagnosis, duration of symptoms before diagnosis, presence of additional autoimmune conditions, and injection frequency. Odds ratios were reported with 95% confidence intervals.

Full methodological details are provided in the Supplementary Methods (**Supplementary Notes 2-5**), including descriptive analyses, injection-cycle construction, individualised response-window modelling, and responder phenotyping with multilevel clustering analyses.

### Construction of Symptom Domains

To examine broader patterns of symptom response beyond individual symptoms, related symptoms were grouped into clinically and biologically informed domains. Three domains were constructed: cognitive (poor concentration, memory loss, nominal aphasia), neuro-sensory (paraesthesia, nerve pain, dizziness, tinnitus), and pain-fatigue-affective (fatigue, myalgia, irritability). Further details on domain construction and rationale are provided in **Supplementary Note 6**.

A composite Global Symptom Burden score was calculated as the mean severity across all recorded symptoms (excluding overall wellbeing), with absent symptoms contributing a score of zero.

### Ethics

The study protocol and procedures were reviewed and given a Favourable Ethical Opinion by the University of Surrey Ethics Committee (EGA ref: FHMS 22-23 030 EGA). All participants provided electronic informed consent before participation

## Results

### Participant characteristics

A total of 86 participants were enrolled in the study. Fifteen participants withdrew during follow-up: four because of work or other commitments, one after starting a new medication whose side effects could interfere with study assessments, and two because of illness; eight did not provide a reason for withdrawal. A further three participants were excluded because of low survey completion (<20%), leaving 68 participants in the longitudinal analyses. **Table 1** summarises the key characteristics of these participants. The sample was predominantly female (82% female, 18% male). The mean age of participants was 63 years (SD=11, range 27-85 years old). The mean age of PA diagnosis was 48 years (SD=14 | range 14-73).

**Table 1.** Demographic and clinical characteristics of participants in the daily symptom-tracking study (n= 68).

| **Characteristic** | **N (%)** |
|---|---|
| **Sex (N, %)** |  |
| Male | 12 (18) |
| Female | 56 (82) |
| **Age (Mean, SD)** |  |
| Age | 63 (11) |
| Age diagnosed | 48 (14) |
| **Length of time with diagnosis (N, %)** |  |
| < 6 months | 8 (12) |
| 6 months - 1 year | 10 (15) |
| 1-3 years | 19 (28) |
| 3-5 years | 10 (15) |
| 5 years + | 21 (31) |
| **Familial Pernicious Anaemia (N, %)** |  |
| Yes | 43 (63) |
| No | 25 (37) |
| **Other autoimmune diseases (N, %)** |  |
| Yes | 40 (59) |
| No | 28 (41) |
| **Treatment Type (N,%)<sup>¥</sup>** |  |
| Sublingual | 270 (30) |
| Intramuscular Injection | 243 (27) |
| Spray | 198 (22) |
| Oral Supplement | 142 (16) |
| Subcutaneous Injection | 42 (5) |
| Patch | 1 (<1) |

<sup>¥</sup> Participants could report multiple vitamin B12 administration methods during the study period; counts therefore represent reported treatment episodes rather than unique participants, and percentages represent the proportion of all reported treatment episodes.

A total of 5,165 daily reports were recorded (84% of 6,131 expected observations). On average, each participant contributed 76 days of data (SD 28), though individual completion rates varied from 21% to 100%.

### Vitamin B_12_ treatment & Injection frequency

At the time of the study, all participants were receiving maintenance B_12_ injections administered as 1000 µg doses with varying durations. The largest subgroup (31%) had been diagnosed with PA for more than five years. **Table 1** provides a breakdown of the different types of B_12_ treatments used by participants. Injections (whether intramuscular or subcutaneous) represented the most common treatment method (n=285, 32%); Standard 2-3 monthly injections were the most common treatment schedule (36/68, 53%) [21]. The non-standard category included: Monthly (18/68, 26%), every 2-3 weeks (6/68, 9%), and weekly or more frequent injections (8/68, 12%).

### Symptom prevalence and severity

**Fig 1** shows the frequency of each symptom reported during the study by all participants. Fatigue was the most commonly reported symptom (59% of all daily entries included Fatigue) and dizziness the least common (22%), with the remaining symptoms reported in 27-40% of daily entries. Symptom severity was generally moderate-to-high across the cohort, with median scores typically ranging between 5-6 out of 10 (IQR≈3 for most symptoms) (**Supplementary Table 2**). Fatigue showed the highest mean severity (6.34±1.97), whereas paraesthesia had the lowest mean severity (4.81±2.40); Standard deviations were generally large (SD∼2), indicating substantial variability in symptom severity.

**Fig 1.**
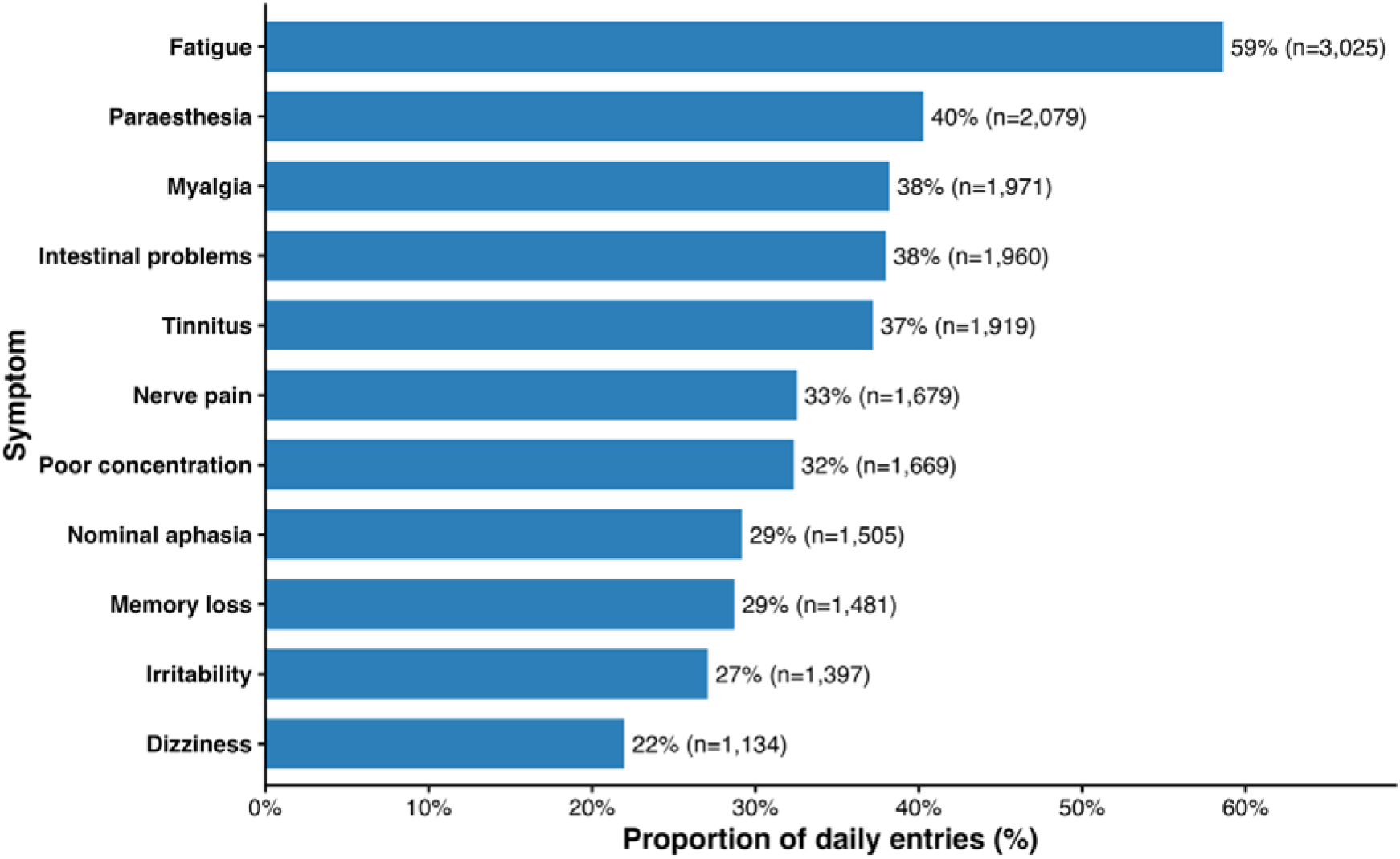
Frequency of symptom reports during daily symptom tracking by 68 adults with Pernicious Anaemia. Bars show the proportion of completed daily entries (participant-days) in which each symptom was reported (score > 0). Percentages are calculated using the total number of completed daily entries as the denominator (n= 5,165). Multiple symptoms could be reported on the same day; therefore, percentages do not sum to 100%.

### Partitioning within- and between individual symptom variability

Inter-symptom clustering was quantified using pooled Spearman rank correlations, and symptom variability was partitioned using ICCs. Symptom correlations ranged from -0.05 to 0.65 (**Supplementary Fig 1**). The strongest correlations were observed among cognitive symptoms, particularly poor concentration with memory loss (r=0.62) and nominal aphasia (r=0.54), suggesting a shared cognitive symptom cluster. Pain-related symptoms also showed moderate correlations e.g. paraesthesia with nerve pain, r=0.49, whereas tinnitus demonstrated consistently weak correlations with all other symptoms (typically r<0.20). ICCs showed that most symptom variability was attributable to stable between-participant differences rather than within-participant fluctuation (0.58-0.80; **Supplementary Table 3**).

### Symptom scores decrease following vitamin B12 injection

Pre- and post-treatment scores were calculated as the median symptom score across the three days immediately before and after each injection. Across all symptoms and composite domains, post-treatment median scores were significantly lower than pre-treatment scores (**Table 2**; all p<0.001). The largest median reductions were observed for fatigue and cognitive symptoms. Fatigue showed the greatest absolute improvement of all symptoms (Pre: 6.33 [IQR 2.56]; Post: 3.86 [IQR 4.99]; Δ -2.47, -39.0%; N=54), followed by poor concentration and memory loss. Neuro-sensory symptoms also showed marked improvements, whereas gastrointestinal symptoms demonstrated more modest improvements.

**Table 2.** Participant-averaged changes in symptom severity and overall wellbeing 3 days pre-versus 3 days post-vitamin B_12_ injection among adults with pernicious anaemia (N=62).

| **Outcome** | **N** | **Pre-Median (IQR)<sup>a</sup>** | **Post-Median (IQR)<sup>b</sup>** | **Δ** | **% Change<sup>c</sup>** | **N injection events** |
|---|---|---|---|---|---|---|
| Dizziness | 26 | 3.42 (3.08) | 1.08 (4.51) | -2.33 | -68.3% | 58 |
| Intestinal problems | 36 | 4.71 (3.89) | 3.5 (5.51) | -1.21 | -25.7% | 102 |
| Irritability | 30 | 3.92 (2.45) | 2.62 (4.35) | -1.29 | -33% | 74 |
| Memory loss | 30 | 4.38 (2.92) | 2.25 (3.59) | -2.12 | -48.6% | 83 |
| Myalgia | 41 | 5.5 (2.83) | 3.67 (3.87) | -1.83 | -33.3% | 127 |
| Nerve pain | 32 | 5.21 (2.62) | 3.96 (3.9) | -1.25 | -24% | 91 |
| Paraesthesia | 36 | 3.88 (3.36) | 2.11 (3.58) | -1.76 | -45.5% | 99 |
| Poor concentration | 38 | 4.62 (2.84) | 2.28 (3.76) | -2.34 | -50.7% | 103 |
| Tinnitus | 32 | 5.05 (3.89) | 3.67 (4.75) | -1.38 | -27.4% | 86 |
| Fatigue | 54 | 6.33 (2.56) | 3.86 (4.99) | -2.47 | -39% | 161 |
| Nominal aphasia | 28 | 4.33 (2.96) | 3.06 (3.33) | -1.28 | -29.5% | 83 |
| Overall wellbeing | 60 | 4.96 (2.56) | 3.62 (3.54) | -1.34 | -26.6% | 187 |
| Composite Symptom Burden | 62 | 4.66 (1.96) | 2.79 (2.4) | -1.87 | -40.2% | 187 |
| Cognitive mean | 44 | 2.59 (2.75) | 1.36 (2.44) | -1.23 | -47.4% | 122 |
| Neuro mean | 55 | 1.75 (2.1) | 1.25 (2.33) | -0.5 | -28.6% | 158 |
| Pain fatigue mean | 59 | 3.51 (2.81) | 2.33 (2.85) | -1.18 | -33.5% | 173 |

Symptom scores were rated on a 0-10 scale (0 = none; 10 = most severe). Values are medians (IQR) of participant-level averages. For each eligible injection, median symptom severity was calculated across the three-day pre- and post-injection windows; where participants contributed multiple eligible injections, these injection-level medians were averaged within participant to avoid over-representation of frequent injectors.Wilcoxon signed-rank test comparing paired participant-level pre- versus post-treatment averages were all highly significant (P<0.001). For symptom-specific analyses, participants contributed values only where the symptom was present during the pre-injection period. Participants who did not report a given symptom during the relevant pre-injection period did not contribute to that symptom-specific analysis; post-injection scores of zero were retained and represented symptom resolution.
<sup>a</sup>Pre-treatment period: median symptom severity across the 3 days immediately prior to injection.
<sup>b</sup>Post-treatment period: median symptom severity across the 3 days immediately following injection.
<sup>c</sup>% change: median-based percentage change calculated as (Median.post – Median.pre) / Median.pre) x 100.
N: number of participants contributing non-missing paired values for that outcome
N injection events: number of eligible isolated injections contributing to that outcome (distinct participant–injection pairs)

Composite domain scores showed a similar pattern, with the cognitive domain demonstrating the greatest reduction (Pre: 2.59 [IQR 2.75]; Post: 1.36 [IQR 2.44]; Δ -1.23, - 47.4%; p < 0.001; N = 44), followed by the pain-fatigue and neuro-sensory domains. Composite symptom burden demonstrated a large and consistent treatment response (Pre: 4.66 [IQR 1.96]; Post: 2.79 [IQR 2.40]; Δ -1.87, -40.2%; p < 0.001; N = 62). Overall wellbeing also improved significantly following treatment (Pre: 4.96 [IQR 2.56]; Post: 3.62 [IQR 3.54]; Δ -1.34, -26.6%; p < 0.001; N = 60).

Sensitivity analyses using individualised response windows (**Supplementary Table 4**) confirmed the direction of the primary 3-day findings while demonstrating larger absolute treatment effects.

To evaluate whether symptom score improvements differed by injection frequency, median changes were compared between standard (2-3 monthly) and non-standard (monthly, every 2-3 weeks, and weekly or more frequent) regimens (**Fig 2**). The magnitude of response varied by symptom and domain rather than following a uniform dose-response pattern (**Supplementary Table 5**). For example, cognitive symptoms demonstrated greatest improvements under standard 2-3 monthly regimens, whereas fatigue, pain-related and several neuro-sensory symptoms showed larger improvements with more frequent dosing schedules. Overall wellbeing improved across all treatment frequency groups, with the largest reduction in the wellbeing score observed among participants receiving non-standard regimens.

**Fig 2.**
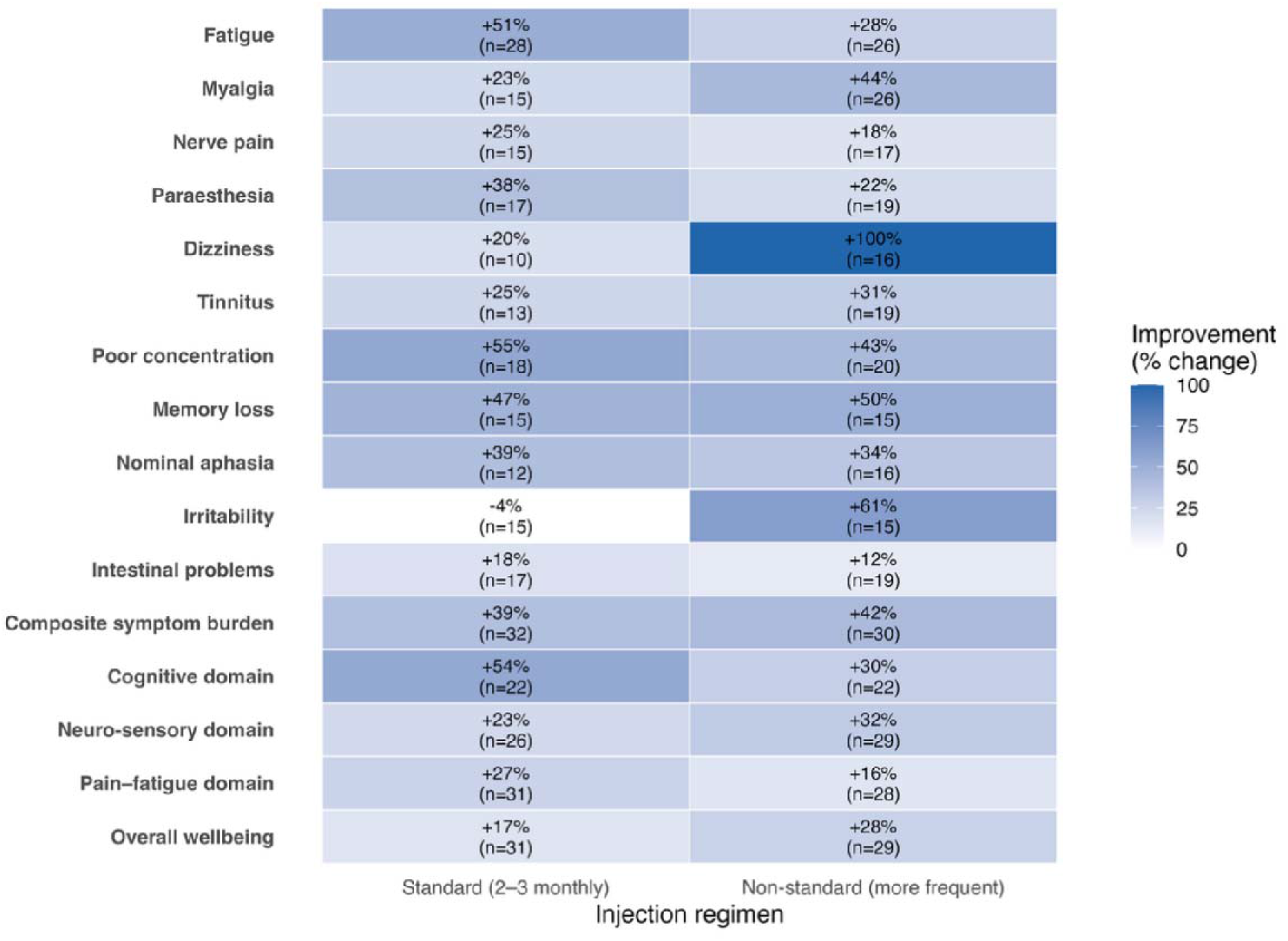
Symptom and overall wellbeing change by vitamin B_12_ injection regimen (Standard vs Non-standard)

Heatmap showing median percentage change in symptom severity and domain-level composite scores following intramuscular or subcutaneous vitamin B12 injections, stratified by predominant injection regimen (standard 2–3 monthly vs non-standard more frequent dosing). Values represent the median percentage change between the 3-day pre-injection and 3-day post-injection windows. For symptom and domain measures, positive values indicate a reduction in symptom severity; for overall wellbeing, positive values indicate improvement in wellbeing. Cell labels show median percentage change and the number of participants contributing paired pre- and post-injection data (n). Colour intensity represents the magnitude of improvement. Differences between regimens were symptom-specific rather than consistently favouring either regimen; for example, dizziness and irritability showed larger improvements under more frequent regimens, whereas the cognitive domain showed a larger improvement under standard 2-3-monthly treatment.

### Temporal dynamics of symptom response to vitamin B_12_ treatment

To visualise temporal symptom dynamics following B_12_ injection, symptom trajectories were aligned to the day of injection (Day 0) and plotted at both the cohort and individual levels (**Fig 3a,b**). Distinct symptom-specific response kinetics were observed. Median symptom scores increased in the days immediately preceding injection and declined sharply thereafter, consistent with injections being administered during periods of rising symptom burden.

**Fig 3.**
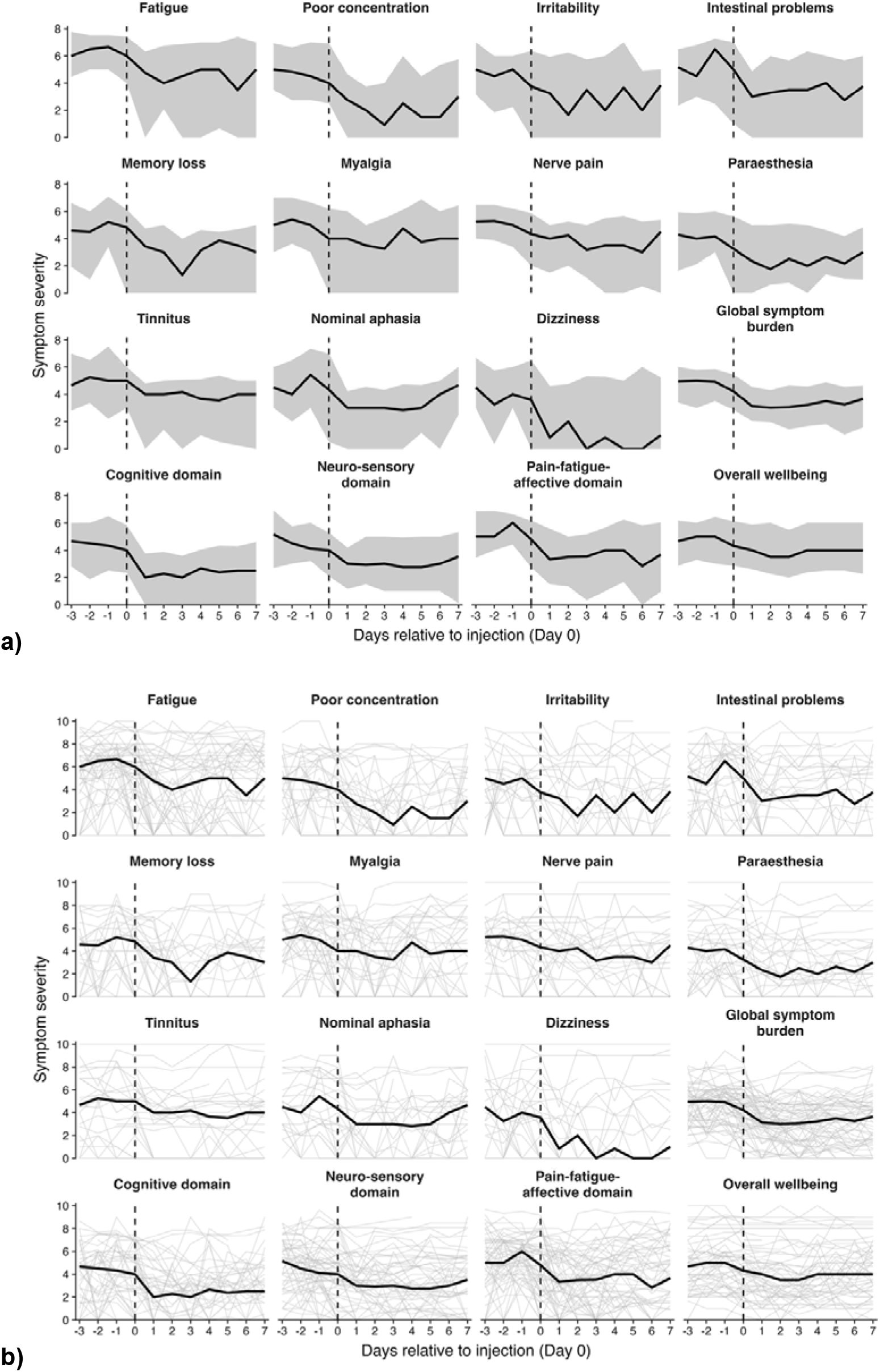
Temporal kinetics of symptom response following vitamin B12 injection. (a) Cohort-level injection-aligned symptom trajectories. Black lines represent median symptom severity and grey shaded areas represent the interquartile range across days relative to injection (Day 0). (b) Individual injection-aligned trajectories (grey), with the cohort median trajectory overlaid in black, illustrating substantial heterogeneity in baseline symptom burden and post-injection response. Together, the panels demonstrate symptom-specific differences in both the timing and variability of response, including rapid early changes in cognitive symptoms and more gradual trajectories for fatigue and pain-related symptom

Cognitive symptoms (poor concentration, memory loss and nominal aphasia) demonstrated rapid early improvement, with median severity falling sharply between Day 0 and Day 1 and reaching a plateau by Days 2-3 before gradual recurrence. In contrast, fatigue and myalgia showed more gradual and sustained improvement, with fatigue reaching its lowest median severity around Day 6 and remaining below pre-treatment levels throughout the observation period. Several symptoms displayed oscillatory (“yo-yo”) patterns across Days 1-7, indicating modal rather than linear recovery trajectories.

In general, neuro-sensory symptoms (paraesthesia, nerve pain, tinnitus, dizziness) showed greater heterogeneity and more delayed response kinetics. Paraesthesia and nerve pain exhibited progressive reductions peaking around Days 2-3, while dizziness demonstrated rapid early improvement with near-complete median resolution by Days 3-4. Composite symptom burden and the cognitive and neuro-sensory domain scores closely mirrored the dominant trajectories of their component symptoms, showing sharp early reductions followed by stabilisation. Visualisation of individual injection trajectories further highlighted marked inter-individual variability in baseline symptom scores and substantial heterogeneity in the magnitude and durability of the post-injection response (**Fig 3b & Supplementary Fig 2**).

Wellbeing scores declined following injection, with the greatest improvement occurring within the first few post-injection days, before remaining generally better than pre-injection levels across the subsequent observation period.

### Differential symptom trajectories in Responders and Non-responders

Participants were stratified by treatment response to compare symptom trajectories between responders and non-responders (**Supplementary Table 6**). Responders demonstrated consistent reductions in symptom scores across individual symptoms and composite domains following B_12_ injections, whereas non-responders showed little or no change and, in some cases, worsening of symptoms (**Supplementary Note 7**; **Supplementary Table 6**).

The participant-by-symptom heatmap (**Fig 4**) showed clear within-participant clustering of treatment response across symptom domains. Participants were grouped according to predominant treatment frequency and ordered by the proportion of symptom responses classified as responders within each treatment-frequency group. Multilevel logistic regression further showed that treatment response was driven predominantly by participant-level rather than symptom-level differences: a participant-level random-intercept model provided the best fit, whereas symptom-level random effects were negligible (ICC=0.008).

**Fig 4.**
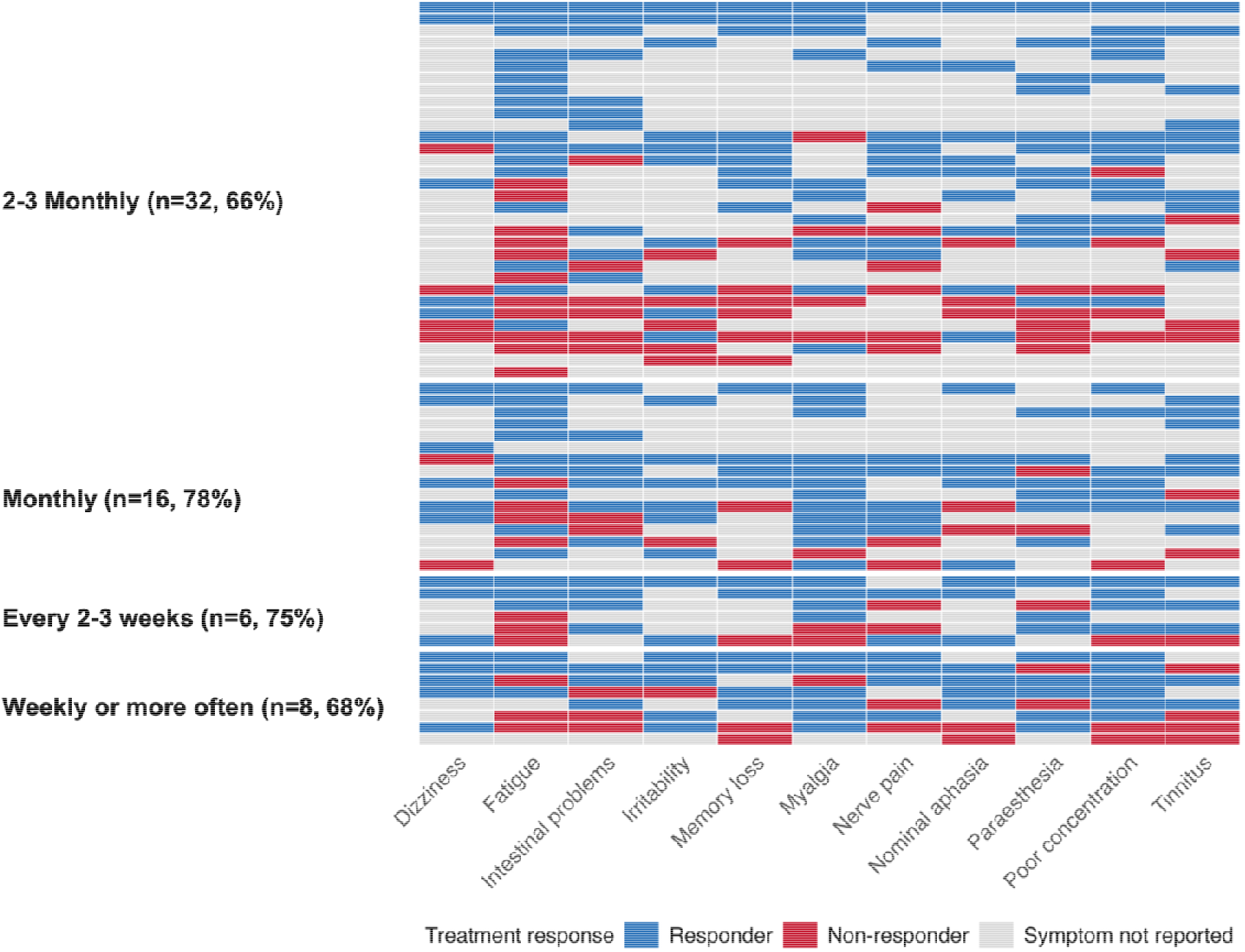
Participant vs symptom heatmap showing clustering of treatment response to vitamin B12 (n=62). Each row represents a participant and each column a symptom. Cells indicate symptom-level treatment-response classification (blue = responder; red = non-responder; grey = symptom not reported). Participants are grouped according to predominant injection frequency (2-3 monthly, monthly, every 2-3 weeks, or weekly or more often) and ordered within each group according to the proportion of symptoms classified as responders. Percentages shown alongside each treatment-frequency group represent the proportion of evaluable participant–symptom responses classified as responders within that group. The heatmap illustrates marked heterogeneity between participants alongside clustering of response within individuals across symptoms.

No demographic or clinical variables were strongly associated with predominant responsiveness; however, more frequent injection showed a non-significant trend toward greater responsiveness (OR 2.37, 95% CI 0.79-7.58; p=0.13). Sensitivity analyses of varying post-injection and baseline window definitions demonstrated stable treatment effect direction and responder classification, supporting the three-day window approach (**Supplementary Note 8**).

## Discussion

This study provides the first longitudinal characterisation of symptom dynamics and treatment response among individuals living with PA using daily symptomatology tracking. Despite all participants receiving long-term maintenance B_12_ therapy, many continued to experience substantial symptom burden with cyclical worsening before injections and rapid improvement following treatment. Across the cohort, symptom severity varied markedly between individuals but was relatively stable within individuals over time, indicating persistent inter-individual symptomatology. In general, B_12_ injections produced rapid and clinically meaningful symptom improvements across multiple domains, although the magnitude, timing, and durability of response varied substantially both between symptoms and between individuals. Notably, treatment responses clustered strongly within individuals across symptom systems, revealing a latent responder/non-responder phenotype that transcends individual symptom categories.

The present study is consistent with prior literature showing that PA has a broad and heterogeneous symptom profile spanning cognitive, neurological and systemic domains [3], [7], [10], [14], [22]. It extends the literature by quantifying within-person symptom dynamics and injection-aligned response kinetics. One prior study has quantitatively examined patient-reported symptom response to B_12_ treatment at scale. Kornic et al. (2016) conducted a retrospective survey assessing historical treatment regimen, demonstrating that injection therapy and more frequent dosing were associated with greater symptom improvement than oral therapy and long-interval maintenance schedules [23]. While conceptually in line with our findings, their analyses were based on long-term recall and treatment averages and could not characterise temporal kinetics or individual response phenotypes.

The findings also support previous suggestions that symptom improvement may begin within days but can evolve over weeks to months, particularly for neurological symptoms [9], [15], [24], [25]. However, the timing and duration of improvement have not previously been quantified at high temporal resolution or at the level of individual symptoms and injection events.

### PA is a Heterogeneous Symptomatic Condition Warranting Precision Management

Daily symptom tracking demonstrated that symptom burden in PA is characterised stable person-specific symptoms across cognitive, neurological and systemic domains. Neuro-sensory symptoms such as paraesthesia and dizziness exhibited the highest between-person stability, whereas cognitive symptoms showed greater within-person fluctuation. These patterns may reflect differences in the mechanisms underlying symptom persistence and responsiveness. Correlations among cognitive symptoms further demonstrated symptom clustering, suggesting that some symptoms vary together rather than independently.

Treatment response also clustered strongly within participants across symptoms and symptom domains, indicating that individuals with PA are not likely to follow a uniform therapeutic response. Instead, participants exhibit distinct treatment-response phenotypes with varying kinetic profiles and treatment sensitivities. These findings suggest that fixed-interval replacement schedules may not capture the range of treatment responses observed in practice. While observational and not causal, the divergence in symptom trajectories across individuals supports further evaluation of more flexible and pragmatic dosing strategies.

### Delayed and Multisystem Response Kinetics Reveal Latent Responder Phenotypes

A central finding of this study is that response to B_12_ injections is not only heterogeneous in magnitude but also in temporal kinetics, revealing distinct response kinetics across participants. Across symptoms, improvement did not occur uniformly in the immediate post-injection period. Cognitive symptoms exhibit rapid responses, typically peaking within 24-72 hours, whereas fatigue and pain-related symptoms showed more gradual but sustained improvement, and neuro-sensory symptoms exhibited delayed, heterogeneous, and often oscillatory (“yo-yo”) response trajectories.

Importantly, treatment responses clustered within individuals across symptoms and domains, with participants often showing broadly concordant improvement or minimal change across multiple symptom groups. However, as symptom severity was self-reported, this pattern could also partly reflect consistent reporting behaviour or expectancy effects, whereby individuals’ beliefs about treatment effectiveness influence symptom ratings across domains. Given the observational design, our findings cannot distinguish biological treatment sensitivity from reporting-related effects. Rather, they indicate stable individual-level response patterns that warrant further investigation under controlled conditions.

### Injection Frequency and Symptom-Specific Treatment Response

Injection frequency did not exert a uniform effect on symptom burden across the cohort. Importantly, these findings were observed among individuals receiving long-term maintenance B_12_ therapy, who would generally be expected to have stable symptom control. Instead, many participants continued to experience recurrent symptom exacerbations between injections, suggesting that maintenance treatment did not consistently provide sustained symptom control.

Distinct symptom- and domain-specific dose-response patterns were observed, indicating that optimal injection frequency may vary by symptom system rather than following a simple “more is better” paradigm. Notably, the highest injection frequencies were not associated with greater multisystem symptom control; composite symptom burden showed its largest improvement under monthly regimens, whereas weekly-or-more-frequent dosing was associated with comparatively little improvement. However, these findings should be interpreted cautiously, as reported injection frequency may reflect attempts to improve symptom control rather than objectively an individual’s optimal treatment regimen.

Frequent injection use may reflect two distinct clinical scenarios. In some individuals, rapid symptom recurrence despite preserved responsiveness may indicate undertreatment under standard maintenance schedules. In others, escalating injection frequency may represent a compensatory response to reduced responsiveness to B_12_ treatment, whereby increasing injection frequency provides limited symptomatic benefit. The relatively modest treatment effects observed in this group, together with strong within-person clustering of treatment response across symptom domains, suggest that persistent symptoms may not be explained solely by insufficient dosing. Alternative explanations include reduced biological responsiveness to B_12_ treatment, irreversible neurological sequelae following delayed diagnosis, or symptom recurrence after transition from more frequent to standard maintenance regimens [26], [27], [28]. This may help explain why increasing injection frequency does not uniformly restore symptom control in all participants. Collectively, these findings support a more individualised, symptom-guided approach to maintenance treatment rather than a single fixed dosing schedule.

Finally, almost half of participants reported receiving B_12_ injections more frequently than the current standard 2-3-month maintenance regimen, and many also used additional over-the-counter B12 preparations e.g. oral, sublingual between injections. Although adjunctive therapies were not formally evaluated, their widespread use may reflect unmet symptom control needs within current maintenance schedules. This may suggest that some individuals do not achieve satisfactory symptom relief with any fixed-interval replacement alone [15], [21], [29], although it may also suggest adherence to a more is better approach.

## Strengths and Limitations

This study represents the first high-resolution longitudinal characterisation of symptom dynamics and treatment response in PA using daily digital patient-reported outcome measures. Rich within-person sampling and injection-aligned modelling enabled evaluation of symptom trajectories and treatment response beyond what is possible using cross-sectional or recall-based study designs.

Participants demonstrated high engagement with daily symptom tracking, with an overall completion rate of 84% and a mean of 76 days of data contributed per participant (SD 28). direct comparison with other remote monitoring studies is limited by inconsistent reporting of adherence [30], this supports the feasibility and scalability of longitudinal digital symptom tracking in PA.

Several limitations should be acknowledged. Recruitment through a patient organisation may have preferentially included individuals with greater symptom burden or engagement with their condition, potentially limiting generalisability to the wider PA population. Symptom severity and treatment exposure were self-reported, introducing potential reporting bias and misclassification. Although daily prospective reporting substantially reduces recall bias compared with retrospective surveys, symptom ratings remain subjective and may be influenced by symptom awareness or expectations of treatment response [31]. Although high-frequency sampling improves temporal resolution, individual response kinetics may extend beyond the post-injection windows captured in this study. Consequently, a subset of participants classified as non-responders may include delayed or atypical responders whose symptom improvements fall outside the analysed response periods. In addition, co-existing nutritional deficiencies (e.g. iron and/or folate deficiency) were not evaluated and may also have contributed to symptom burden. Although the within-person analytic design mitigates many time-invariant confounders, residual confounding from non-PA-related symptom drivers cannot be excluded.

## Future Work

This work establishes a foundation for precision-guided management of PA and highlights several priorities for future research. First, mechanistic studies are needed to define the biological determinants underlying stable responder phenotypes and symptom-specific treatment kinetics. Understanding the drivers of differential responsiveness will be essential for explaining refractory symptom patterns and informing targeted therapeutic strategies.

Second, intervention studies comparing fixed-interval dosing with personalised injection schedules tailored to individual symptom trajectories are warranted to establish evidence-based maintenance models for PA. Although the present study cannot determine causality or optimal dosing strategies, the observed heterogeneity in treatment response provides a strong rationale for pragmatic trials evaluating symptom-guided maintenance approaches.

Third, integration of longitudinal digital symptom monitoring into clinical care requires formal evaluation. Digital platforms enabling real-time symptom tracking, trajectory visualisation and clinician-linked data sharing may support more individualised treatment adjustment. Such approaches may also complement conventional biomarker monitoring by providing additional information on day-to-day symptom burden and treatment response. Preliminary pilot work using wearable smartwatch technology (unpublished data) has demonstrated the feasibility of continuous physiological monitoring in PA, supporting future integration of objective digital measures alongside symptom-based tracking within precision treatment models.

## Conclusions

PA is a heterogeneous multisystem condition with marked variability in symptom burden and treatment responsiveness despite maintenance B_12_ therapy. B_12_ injections produced rapid symptom improvement, but response kinetics varied substantially between and within individuals and symptom domains. These findings support evaluation of more individualised, symptom-guided maintenance strategies.

## Data Availability

The data supporting the findings of this study are not publicly available but are available from the corresponding author upon request.

## Acknowledgments

The authors thank Professor Bruce H.R. Wolffenbuttel and Professor Ebba Nexø for their valuable comments and feedback on the work. We also thank the Pernicious Anaemia Society and its members for their support of the study and all participants who generously contributed their time and daily symptom data.

## References

[1] B.-H. Toh, I. R. van Driel, and P. A. Gleeson, “Pernicious Anemia,” New England Journal of Medicine, vol. 337, no. 20, pp. 1441–1448, Nov. 1997, doi: 10.1056/NEJM199711133372007.

[2] A. Hunt, D. Harrington, and S. Robinson, “Vitamin B12 deficiency,” Sep. 04, 2014, BMJ Publishing Group. doi: 10.1136/bmj.g5226.

[3] M. Hooper, P. Hudson, F. Porter, and A. McCaddon, “Patient journeys: diagnosis and treatment of pernicious anaemia,” British Journal of Nursing, vol. 23, no. 7, pp. 376–381, Apr. 2014, doi: 10.12968/bjon.2014.23.7.376.

[4] A. Thain et al., “Patient-reported characteristics of pernicious anaemia: a first step to initiate James Lind Alliance Priority Setting Partnership driven research,” BMC Primary Care, vol. 26, no. 1, p. 348, Nov. 2025, doi: 10.1186/s12875-025-03036-0.

[5] K. Staley et al., “Research priorities in pernicious anaemia: James Lind Alliance Priority Setting Partnership,” BMJ Open, vol. 12, no. 8, p. e065166, Aug. 2022, doi: 10.1136/bmjopen-2022-065166.

[6] N. Tyler, A. Hodkinson, N. Ahlam, S. Giles, A. Zhou, and M. Panagioti, “Patient safety, self-injection, and B12 deficiency: a UK cross-sectional survey,” British Journal of General Practice, vol. 72, no. 725, pp. e891–e898, Dec. 2022, doi: 10.3399/BJGP.2021.0711.

[7] E. B. Healton, D. G. Savage, J. C. M. Brust, T. J. Garrett, and J. Lindenbaum, “Neurologic Aspects of Cobalamin Deficiency,” Medicine, vol. 70, no. 4, pp. 229–245, Jul. 1991, doi: 10.1097/00005792-199107000-00001.

[8] D. G. Savage and J. Lindenbaum, “11 Neurological complications of acquired cobalamin deficiency: clinical aspects,” Baillieres Clin. Haematol., vol. 8, no. 3, pp. 657–678, Sep. 1995, doi: 10.1016/S0950-3536(05)80225-2.

[9] J. Lindenbaum et al., “Neuropsychiatric Disorders Caused by Cobalamin Deficiency in the Absence of Anemia or Macrocytosis,” New England Journal of Medicine, vol. 318, no. 26, pp. 1720–1728, Jun. 1988, doi: 10.1056/NEJM198806303182604.

[10] C. H. Seage et al., “A Systematic Review of Symptoms of Pernicious Anemia,” Food Nutr. Bull., vol. 45, no. 1_suppl, pp. S34–S39, Jun. 2024, doi: 10.1177/03795721241227016.

[11] R. Green et al., “Vitamin B12 deficiency,” Nat. Rev. Dis. Primers, vol. 3, Jun. 2017, doi: 10.1038/nrdp.2017.40.

[12] S. N. van der Veer et al., “Remote monitoring of rheumatoid arthritis (REMORA): study protocol for a stepped wedge cluster randomized trial and process evaluation of an integrated symptom tracking intervention.,” Trials, vol. 25, no. 1, p. 683, Oct. 2024, doi: 10.1186/s13063-024-08497-9.

[13] J. Gandrup, D. A. Selby, S. N. van der Veer, J. Mcbeth, and W. G. Dixon, “Using patient-reported data from a smartphone app to capture and characterize real-time patient-reported flares in rheumatoid arthritis,” Rheumatol. Adv. Pract., vol. 6, no. 1, Jan. 2022, doi: 10.1093/rap/rkac021.

[14] C. Briani et al., “Cobalamin Deficiency: Clinical Picture and Radiological Findings,” Nutrients, vol. 5, no. 11, pp. 4521–4539, Nov. 2013, doi: 10.3390/nu5114521.

[15] National Institute for Health and Care Excellence. Vitamin B12 deficiency in over 16s: diagnosis and management. NICE guideline NG239. London: NICE; 2024. Available from: https://www.nice.org.uk/guidance/ng23916.

[16] J. T. Farrar, J. P. Young, L. LaMoreaux, J. L. Werth, and M. R. Poole, “Clinical importance of changes in chronic pain intensity measured on an 11-point numerical pain rating scale,” Pain, vol. 94, no. 2, pp. 149–158, Nov. 2001, doi: 10.1016/S0304-3959(01)00349-9.

[17] R. W. J. G. Ostelo et al., “Interpreting Change Scores for Pain and Functional Status in Low Back Pain,” Spine (Phila. Pa. 1976)., vol. 33, no. 1, pp. 90–94, Jan. 2008, doi: 10.1097/BRS.0b013e31815e3a10.

[18] F. Salaffi, A. Stancati, C. A. Silvestri, A. Ciapetti, and W. Grassi, “Minimal clinically important changes in chronic musculoskeletal pain intensity measured on a numerical rating scale,” European Journal of Pain, vol. 8, no. 4, pp. 283–291, Aug. 2004, doi: 10.1016/j.ejpain.2003.09.004.

[19] G. H. Guyatt, E. F. Juniper, S. D. Walter, L. E. Griffith, and R. S. Goldstein, “Interpreting treatment effects in randomised trials,” BMJ, vol. 316, no. 7132, pp. 690–693, Feb. 1998, doi: 10.1136/bmj.316.7132.690.

[20] R. H. Dworkin et al., “Interpreting the Clinical Importance of Treatment Outcomes in Chronic Pain Clinical Trials: IMMPACT Recommendations,” J. Pain, vol. 9, no. 2, pp. 105–121, Feb. 2008, doi: 10.1016/j.jpain.2007.09.005.

[21] Joint Formulary Committee. British National Formulary: hydroxocobalamin [Internet]. London: BMJ Group and Pharmaceutical Press; Available from: https://bnf.nice.org.uk/drugs/hydroxocobalamin/

[22] B. H. R. Wolffenbuttel, H. J. C. M. Wouters, M. R. Heiner-Fokkema, and M. M. van der Klauw, “The Many Faces of Cobalamin (Vitamin B12) Deficiency,” Mayo Clin. Proc. Innov. Qual. Outcomes, vol. 3, no. 2, pp. 200–214, Jun. 2019, doi: 10.1016/j.mayocpiqo.2019.03.002.

[23] P. Kornic, M. Harty, and J. Grant, “Influence of Treatment Parameters on Symptom Relief in Individuals with Vitamin B12 Deficiency,” Annu. Res. Rev. Biol., vol. 11, no. 5, pp. 1–8, Jan. 2016, doi: 10.9734/ARRB/2016/31711.

[24] R. Carmel, “How I treat cobalamin (vitamin B12) deficiency,” Blood, vol. 112, no. 6, pp. 2214–2221, Sep. 2008, doi: 10.1182/blood-2008-03-040253.

[25] R. Obeid et al., “Diagnosis, Treatment and Long-Term Management of Vitamin B12 Deficiency in Adults: A Delphi Expert Consensus,” J. Clin. Med., vol. 13, no. 8, p. 2176, Apr. 2024, doi: 10.3390/jcm13082176.

[26] N. Bizzaro and A. Antico, “Diagnosis and classification of pernicious anemia,” Autoimmun. Rev., vol. 13, no. 4–5, pp. 565–568, Apr. 2014, doi: 10.1016/j.autrev.2014.01.042.

[27] B. Hemmer, F. X. Glocker, M. Schumacher, G. Deuschl, and C. H. Lucking, “Subacute combined degeneration: clinical, electrophysiological, and magnetic resonance imaging findings,” J. Neurol. Neurosurg. Psychiatry, vol. 65, no. 6, pp. 822–827, Dec. 1998, doi: 10.1136/jnnp.65.6.822.

[28] A. McCaddon, “Vitamin B12 in neurology and ageing; Clinical and genetic aspects,” Biochimie, vol. 95, no. 5, pp. 1066–1076, May 2013, doi: 10.1016/j.biochi.2012.11.017.

[29] A. Thain, K. Hart, and K. R. Ahmadi, “Addressing the Gaps in the Vitamin B12 Deficiency 2024 NICE Guidelines: Highlighting the Need for Better Recognition, Diagnosis, and Management of Pernicious Anaemia,” Eur. J. Clin. Nutr., Feb. 2025, doi: 10.1038/s41430-025-01583-4.

[30] K. M. White et al., “A systematic review of engagement reporting in remote measurement studies for health symptom tracking,” NPJ Digit. Med., vol. 5, no. 1, p. 82, Jun. 2022, doi: 10.1038/s41746-022-00624-7.

[31] K. MacKrill, K. M. Groom, and K. J. Petrie, “The effect of symptom tracking apps on symptom reporting,” Br. J. Health Psychol., vol. 25, no. 4, pp. 1074–1085, Nov. 2020, doi: 10.1111/bjhp.12459.

