## Supplementary Material for "Symptom variability and treatment response in pernicious anaemia: a longitudinal daily symptom tracking study"

Alfie Thain et al.

**Supplementary Contents**

| **Section** | **Description** | **Page** |
| --- | --- | --- |
| Supplementary Note 1 | Rationale for symptom selection | 2 |
| Supplementary Note 2 | Descriptive analyses | 3 |
| Supplementary Note 3 | Pre-post injection cycle modelling | 4 |
| Supplementary Note 4 | Individualised treatment dynamics | 5 |
| Supplementary Note 5 | Person-level clustering and responder phenotyping | 6 |
| Supplementary Note 6 | Construction of symptom domains | 7 |
| Supplementary Note 7 | Symptom-specific treatment response phenotypes | 8 |
| Supplementary Note 8 | Sensitivity analyses | 9 |
| Supplementary Figure 1 | Symptom correlation matrix | 10 |
| Supplementary Figure 2 | Example participant symptom trajectories | 11 |
| Supplementary Table 1 | Daily survey questions | 12 |
| Supplementary Table 2 | Symptom severity summary statistics | 13 |
| Supplementary Table 3 | Intraclass correlation coefficients | 13 |
| Supplementary Table 4 | Individualised treatment responses | 14 |
| Supplementary Table 5 | Injection frequency analyses | 15 |
| Supplementary Table 6 | Symptom-specific treatment-response classifications | 17 |

**Supplementary Note 1. Rationale for Symptom Selection**

Symptoms were selected a priori based on their prevalence in a large PAS patient survey (3). To ensure inclusion of symptoms most representative of the condition, only symptoms reported by ≥50% of respondents in the previous survey were prioritised.

Based on this criterion, the following symptoms were included: fatigue; memory loss; poor concentration; pins and needles; dizziness; nerve pain; nominal aphasia; myalgia; irritability; intestinal problems; and tinnitus. Intestinal problems were grouped to capture common gastrointestinal manifestations reported in the PAS survey e.g. diarrhoea and indigestion.

Symptom selection was co-developed with individuals living with PA and refined during pilot testing to ensure face validity and relevance to lived experience. A free-text “other symptoms” option was included in the survey instrument to allow reporting of additional symptoms; however, these were not included in the present analyses due to low and heterogeneous reporting frequency.

**Supplementary Note 2. Descriptive analyses**

Baseline symptom burden and injection patterns were summarised prior to treatment-effect modelling. Participants were grouped according to predominant injection frequency (2-3 monthly, monthly, every 2-3 weekly, weekly or more often). These categories reflected the injection frequencies reported by participants and included both standard NHS maintenance regimens and more frequent schedules, including self-administered treatment. For each symptom, median, interquartile range (IQR), mean, and standard deviation (SD) were calculated within frequency groups. Distributional properties, including skewness and kurtosis, were examined to inform the use of non-parametric analytic approaches.

**Supplementary Note 3. Pre-Post Injection Cycle Modelling**

To estimate within-person treatment effects while minimising contamination from adjacent doses, we implemented an injection-cycle modelling framework using temporally aligned daily symptom data.

Daily treatment records were used to identify all intramuscular and subcutaneous vitamin B_12_ injections reported by each participant. An injection occurring on day t was considered eligible for analysis only if it could be isolated from neighbouring injections: specifically, no injection could occur within the preceding three days (t-3 to t-1) and no further injection could occur within the subsequent three days (t+1 to t+3). This ensured that symptom changes attributed to an injection were not influenced by residual effects of adjacent doses.

For each eligible injection cycle, symptom severity was summarised as the mean daily severity across a three-day pre-treatment baseline window (t-3 to t-1) and a three-day post-treatment response window (t+1 to t+3). Injection cycles were retained only if at least one symptom observation was available in both windows.

To avoid dilution of treatment effects by symptom-free baseline periods, injection cycles were excluded for a given symptom if that symptom was absent throughout the pre-treatment window. Post-treatment symptom scores were permitted to be zero, allowing full capture of symptom resolution. Symptoms absent throughout the pre-injection window but reported after treatment were excluded from the primary paired analysis but retained separately as a post-treatment-only category in responder phenotype analyses.

To prevent over-representation of individuals who injected more frequently, injection-level pre- and post-treatment means were first aggregated within each participant. For each symptom and composite domain, participant-level mean pre- and post-treatment severities were then used as the analytic unit for all subsequent cohort-level summaries.

**Supplementary Note 4. Individualised treatment dynamics and symptom structure**

To characterise temporal heterogeneity in post-injection symptom response, a flexible response-window framework was implemented. For each eligible injection cycle, the post-treatment response period was permitted to begin up to five days following injection if symptom severity decreased by at least one point relative to baseline. The response period terminated either when symptom severity increased by two points from the minimum reached or when seven days had elapsed, whichever occurred first. This framework enabled estimation of personalised response kinetics. Individual-level response trajectories were visualised using aligned line plots, with cohort-level trends summarised using rolling median curves.

**Supplementary Note 5. Person-level clustering and responder phenotyping**

To evaluate whether treatment response represented a stable person-level phenotype rather than symptom-specific effects, responder classifications were analysed across participants and symptoms. For each participant, the proportion of symptoms classified as responders was calculated. Participants were categorised as predominantly responsive if >50% of their reported symptoms met responder criteria, and predominantly non-responsive otherwise.

Within-person clustering of response across symptom systems was evaluated using pairwise concordance metrics. For each pair of symptoms, odds ratios and Cohen’s k statistics were calculated to quantify agreement in responder status beyond chance.

To determine whether observed between-participant variability in responder proportions exceeded that expected under random allocation, permutation testing was performed. Responder labels were randomly reassigned within each symptom while preserving overall responder prevalence, and participant-level responder proportions were recalculated. This procedure was repeated across multiple iterations to generate a null distribution against which observed clustering was compared.

To formally partition variance attributable to participant-level versus symptom-level effects, multilevel logistic regression models were fitted with responder status as the binary outcome. Random intercepts were specified for participants and symptoms. Intraclass correlation coefficients (ICC) were derived from model variance components to quantify the proportion of total variance attributable to stable person-level clustering of treatment response.

#### **Supplementary Note 6. Construction of Symptom Domains**

To examine broader patterns of symptom response, related symptoms were grouped into three clinically informed exploratory domains based on symptom characteristics and prior literature on manifestations of vitamin B_12_ deficiency.

**Cognitive Domain** (Poor concentration, memory loss, nominal aphasia)

The cognitive domain was constructed to capture higher-order cortical and subcortical cognitive dysfunction associated with B_12_ deficiency (22,23). Cognitive impairment is a well-recognised neuropsychiatric manifestation of PA and reflects impairment of executive function, working memory, attention, and language processing. These symptoms are strongly linked to demyelination, disrupted methylation pathways, and impaired monoaminergic neurotransmission within fronto-striatal and hippocampal circuits (24,25).

**Neuro-Sensory Domain** (Paraesthesia, nerve pain, dizziness, tinnitus)

The neuro-sensory domain reflects sensory and peripheral nervous system dysfunction arising from disruption of both central and peripheral neural pathways. Paraesthesia and nerve pain represent classic manifestations of peripheral neuropathy in vitamin B12 deficiency, reflecting demyelination and axonal injury within sensory nerve fibres (26). Dizziness and tinnitus were included within this domain as sensory-vestibular manifestations linked to impaired central sensory integration, autonomic dysfunction, and altered auditory-vestibular pathway signalling (27–29). These symptoms are frequently co-reported in pernicious anaemia and share overlapping neurobiological mechanisms involving impaired myelin maintenance, disrupted neurotransmitter synthesis, and neuroinflammatory processes affecting sensory processing networks (22,30,31).

**Pain-Fatigue-Affective Domain** (Fatigue, muscle pain, irritability)

The pain-fatigue-affective domain was constructed to reflect systemic neuroimmune and neurochemical disturbances characteristic of PA and B12 deficiency (25,32,33). Irritability was not conceptualised as a cognitive symptom, but rather as an affective and sickness-behaviour manifestation reflecting central monoaminergic disruption, limbic system dysregulation, neuroinflammation, and fatigue burden (34). These mechanisms cluster biologically with fatigue, muscle pain, malaise, and mood lability, which frequently co-occur and represent a unified systemic symptom phenotype.

**Supplementary Note 7. Symptom-Specific treatment response phenotypes**

To characterise heterogeneity in symptom response beyond average pre-post changes, participants were classified into symptom-specific treatment response categories based on combined absolute and percentage change thresholds (**Supplementary Table 6**).

Cognitive symptoms showed the most consistently favourable response profiles. For poor concentration, 18/41 participants (43.9%) were classified as substantial responders, while only 7/41 (17.1%) were non-responders. Nominal aphasia showed a similarly favourable pattern, with 13/29 participants (44.8%) exhibiting substantial improvement and relatively few classified as non-responders (6/29, 20.7%) or worsened (1/29, 3.4%). Memory loss also responded strongly, with 14/33 participants (42.4%) showing substantial improvement, although a sizeable non-responder group remained (9/33, 27.3%).

Fatigue- and pain-related symptoms displayed substantially greater heterogeneity. For fatigue, 20/55 participants (36.4%) showed substantial improvement, while 9/55 (16.4%) demonstrated only minimal improvement, 18/55 (32.7%) were classified as non-responders, and 3/55 (5.5%) worsened. Myalgia showed a bimodal pattern, with 15/42 participants (35.7%) classified as substantial responders and an equal number (15/42, 35.7%) exhibiting only minimal improvement, alongside non-responders (4/42, 9.5%) and worsening (4/42, 9.5%).

Neuro-sensory symptoms demonstrated marked inter-individual variability. Dizziness showed the highest proportion of substantial responders, with 16/34 participants (47.1%) classified as substantially improved, but also included a notable group who developed symptoms only after treatment (8/34, 23.5%). Paraesthesia and nerve pain showed similarly heterogeneous profiles: for paraesthesia, 12/38 participants (31.6%) were substantial responders while 8/38 (21.1%) were non-responders; for nerve pain, substantial responders (8/34, 23.5%) coexisted with a sizeable non-responder group (9/34, 26.5%), alongside worsening (3/34, 8.8%).

Domain-level analyses reinforced these patterns. The cognitive domain demonstrated the most favourable profile, with 23/48 participants (47.9%) classified as substantial responders and relatively few non-responders (8/48, 16.7%). In contrast, the neuro-sensory domain showed broader dispersion, with substantial (12/58, 20.7%), moderate (14/58, 24.1%), and minimal responders (11/58, 19.0%) alongside a sizeable non-responder group (16/58, 27.6%). The pain-fatigue-affective domain similarly showed substantial (14/60, 23.3%), moderate (12/60, 20.0%), and minimal responders (14/60, 23.3%) each accounting for approximately one-quarter of participants, and 15/60 (25.0%) classified as non-responders.

Global symptom burden demonstrated more modest responsiveness. Although 18/62 participants (29%) showed substantial improvement and 13/62 (21%) moderate improvement, 13/62 (21.0%) showed only minimal change, 11/62 (17.7%) were classified as non-responders, and 7/62 (11.3%) experienced worsening, indicating that multisystem disease burden is less uniformly responsive than individual symptom domains.

**Supplementary Note 8. Sensitivity Analyses of Injection Window Definitions**

**Post-injection window length**

To evaluate whether treatment effect estimates depended on the post-treatment observation window, the primary analysis was repeated using progressively extended post-injection averaging windows from 1 to 7 days. Peak response timing varied across symptoms. For example, dizziness and intestinal problems showed maximal median reductions at Day 1 (-100% and -36%, respectively), muscle pain and word-finding difficulties peaked at Day 2 (-37% and -48%), while poor concentration and memory loss reached maximal reductions at Day 3 (-51% and -49%). More delayed peaks were observed for tinnitus (Day 5) and tiredness (Day 6; -44%). Despite these timing differences, extending the post-treatment window beyond three days did not materially alter overall cohort-level conclusions. Composite symptom burden demonstrated substantial reduction by Day 1 (-28%) with minimal systematic gain beyond Day 3. These findings indicate that while peak response timing varies modestly by symptom, the three-day window captures the dominant early treatment response across outcomes.

**Temporal stability of responder classification**

Responder status was recalculated across post-injection windows of 1-7 days. The absolute number of responders generally peaked at the three-day window. For example, global symptom burden response was observed in 42/62 participants (67.7%) at Day 3 compared with 35/53 (66.0%) at Day 6, reflecting declining eligible sample sizes rather than emergence of additional late responders. Although selected outcomes showed later peaks in responder proportion (e.g., tiredness Day 6: 53.8%; nerve pain Day 5: 64.5%), these occurred within progressively smaller analytic subsets and did not substantively change participant-level classification patterns.

**Sensitivity of baseline definition**

To assess the impact of baseline definition, the pre-injection averaging window was varied from 1 to 7 days while maintaining a fixed three-day post-treatment window. Median symptom severity was highest within the final 1-3 days prior to injection and changed minimally when extending the baseline beyond three days. Longer baselines modestly attenuated pre-post differences by including lower-severity days further from injection, but did not alter the direction or statistical significance of treatment effects.

Supplementary Figure 2 illustrates the symptom trajectories of a single participant across all 11 symptoms, selected because they had complete data and exhibited a broad spectrum of symptom types, allowing clear visualisation of cross-symptom temporal patterns. Poor concentration, fatigue, and irritability show marked post-injection reductions, with repeated intervals of low or minimal symptom severity following treatment. In contrast, memory loss and intestinal problems demonstrate smaller and more variable post-injection changes.

The trajectories display a cyclical pattern in which symptoms decline sharply following each injection and subsequently increase again prior to the next injection. In this participant, symptom escalation becomes apparent approximately 8-12 days after injection across several symptom domains. Across the wider sample, the timing and magnitude of post-injection rebound varied substantially between individuals.

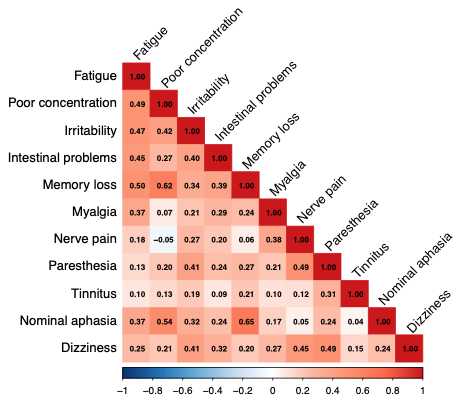

**Supplementary Figure 1.** Symptom correlation matrix of PA symptoms in the daily symptom tracking study (n=68). The matrix illustrates Spearman correlation coefficients between symptom pairs. The values range from -1 to 1, where -1 indicates a strong negative correlation, 0 indicates no correlation, and 1 indicates a strong positive correlation. The colour gradient represents the strength and direction of the correlation: darker red indicates stronger positive correlations, while darker blue represents stronger negative correlations.

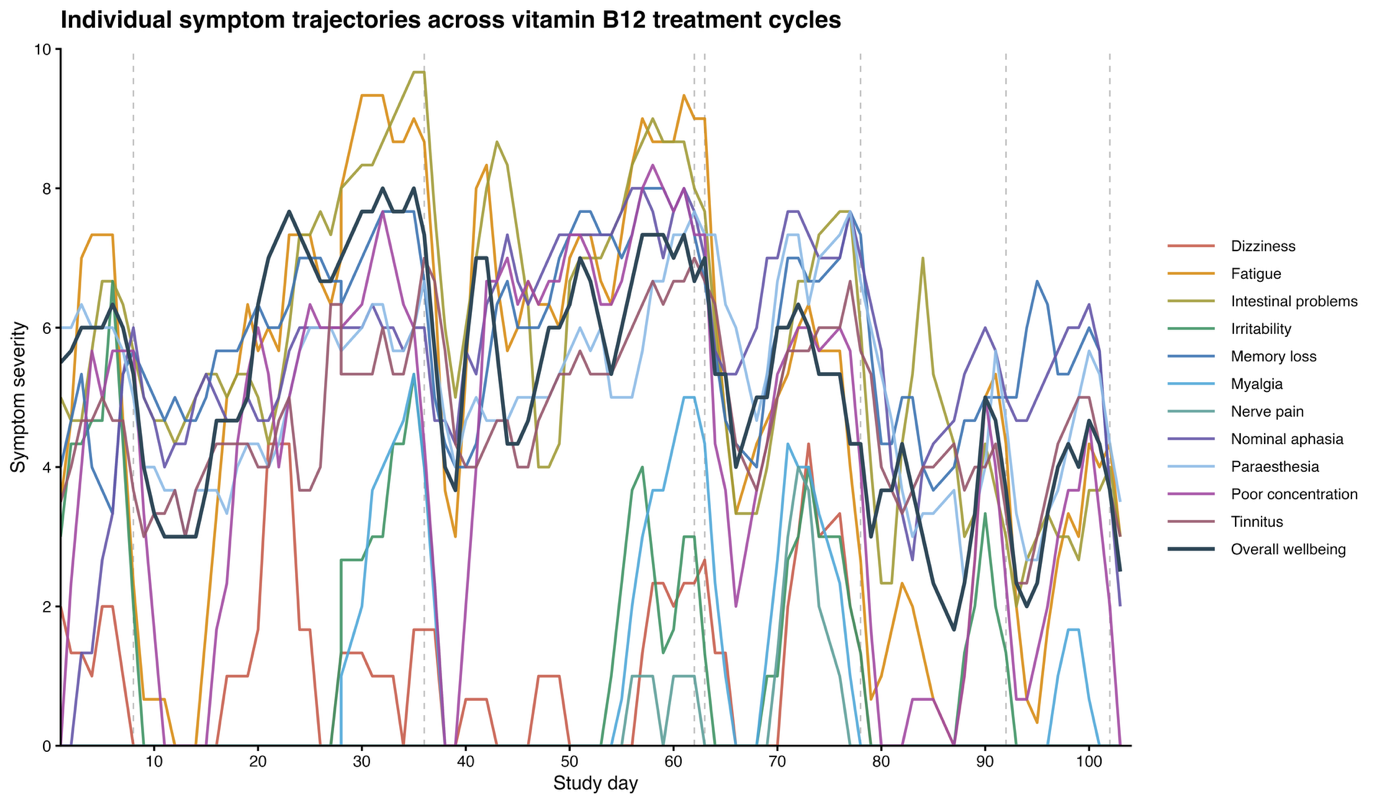

**Supplementary Figure 2.** Individual symptom trajectories from a single participant in the Daily Symptom Tracking Study

Longitudinal symptom trajectories for a representative participant are shown across 11 tracked symptoms. Each coloured line represents the centred 3-day moving average of the daily severity score for an individual symptom, while the dark blue line represents reversed overall self-reported wellbeing (higher values indicate better wellbeing). Dashed vertical lines indicate the timing of vitamin B_12_ injections. Symptom severity was recorded on an 11-point numerical rating scale from 0 (symptom absent) to 10 (most severe).The figure illustrates cyclical symptom dynamics characterised by symptom improvement following vitamin B12 injections and progressive symptom re-emergence before subsequent injections, highlighting within-person temporal patterning and symptom-specific response kinetics.

**Supplementary Table 1.** Overview of daily symptom survey questions.

| **Category** | **Questions** | **Response options** | **Question flow** |
| --- | --- | --- | --- |
| Treatment | 1. Have you had an injection and/or a supplement of vitamin B_12_ today? (e.g. tablets, patches, sublingual) | Yes/no. If yes selected next question if no selected then skip to Q4 | If “Yes,” proceed to Q2; if “No,” skip to Q4. |
|  | 2. What type of vitamin B_12_ have you had? | Set choices: Intramuscular injection, subcutaneous injection, tablets, sublingual, spray, patches | If “Injection” is selected, proceed to Q3; otherwise, skip to Q4. |
|  | 3. What type of vitamin B_12_ injection did you have? Set choices – hydroxocobalamin, cyanocobalamin, methylcobalamin. | Set choices: hydroxocobalamin, cyanocobalamin, methylcobalamin |  |
| Symptomology | 4. Did you experience any symptoms today? Yes/no | Yes/no | If “Yes,” proceed to Q5; if “No,” skip to Q7. |
|  | 5. If yes, What symptoms did you experience today? | List of common PA symptoms with an additional free text option to report other symptoms | Repeat for each reported symptom. |
|  | 6. How severe was X symptom? Please rate it from mild to severe, referring to the symptom definitions. | Scale: 0 (no severity) to 10 (most severe) |  |
| Overall wellbeing | 7. How would you rate your wellbeing on a scale of 1-10 | Scale: 1 (excellent wellbeing) to 10 (poor wellbeing) |  |

**Supplementary Table 2.** Summary of self-reported symptom severity during 3-months daily tracking (n=68).

| **Symptom^a^** | **Mean (SD)** | **Median (IQR)** |
| --- | --- | --- |
| Fatigue | 6.34 (1.97) | 7 (3) |
| Poor Concentration | 5.62 (1.77) | 6 (3) |
| Irritability | 5.7 (2.06) | 6 (3) |
| Intestinal Problems | 5.76 (2.09) | 6 (3) |
| Memory Loss | 5.68 (1.93) | 6 (3) |
| Myalgia | 5.85 (2.06) | 6 (3) |
| Nerve Pain | 5.33 (2.09) | 5 (3) |
| Paraesthesia | 4.81 (2.4) | 5 (3) |
| Tinnitus | 5.25 (2.3) | 5 (3) |
| Nominal aphasia | 5.46 (2.06) | 5 (3) |
| Dizziness | 6.08 (2.47) | 6 (4) |

^a^Symptom severity was rated on a 0-10 numerical rating scale (0 = not present, 10 = most severe). Overall wellbeing was rated from 1 (excellent) to 10 (poor), with lower scores indicating better wellbeing.

**Supplementary Table 3.**  Variance decomposition and intraclass correlation coefficients for daily pernicious anaemia symptoms (n = 68).

| **Variable** | **Variance component** | **Variance** | **ICC*** |
| --- | --- | --- | --- |
| Fatigue | Between participant (subject) | 2.52 | 0.59 |
|  | Residual | 1.75 |  |
| Poor concentration | Between participant (subject) | 1.78 | 0.58 |
|  | Residual | 1.35 |  |
| Irritability | Between participant (subject) | 1.99 | 0.59 |
|  | Residual | 1.38 |  |
| Intestinal problems | Between participant (subject) | 2.49 | 0.59 |
|  | Residual | 1.74 |  |
| Memory loss | Between participant (subject) | 3.1 | 0.69 |
|  | Residual | 1.36 |  |
| Myalgia | Between participant (subject) | 2.60 | 0.61 |
|  | Residual | 1.66 |  |
| Nerve pain | Between participant (subject) | 2.59 | 0.62 |
|  | Residual | 1.59 |  |
| Paraesthesia | Between participant (subject) | 4.09 | 0.80 |
|  | Residual | 1.04 |  |
| Tinnitus | Between participant (subject) | 4.19 | 0.73 |
|  | Residual | 1.55 |  |
| Nominal aphasia | Between participant (subject) | 3.21 | 0.73 |
|  | Residual | 1.19 |  |
| Dizziness | Between participant (subject) | 3.87 | 0.77 |
|  | Residual | 1.18 |  |
| Overall Well-being | Between participant (subject) | 4.67 | 0.78 |
|  | Residual | 1.28 |  |

*The intraclass correlation coefficient (ICC) represents the proportion of total variance attributable to stable between-participant differences. Higher ICC values indicate greater between-participant stability, whereas lower values reflect greater within-participant fluctuation over time.

**Supplementary Table 4.** Summary of individualised treatment responses across symptoms (n=62).

| **Symptom (N)** | **Pre Median**  **(IQR)** | **Post Median**  **(IQR)** | **% Difference^a^** | **Mean Response Duration**  **(Days)^b^** |
| --- | --- | --- | --- | --- |
| Dizziness (20)^c^ | 3.5 (1.25) | 0 (1) | 54.60 | 3.65 |
| Intestinal problems (29) | 5 (1) | 0 (0.5) | 49.58 | 3.79 |
| Irritability (39) | 4 (2.5) | 0 (0) | 68.13 | 4.00 |
| Memory loss (40) | 6 (0.5) | 4 (1) | 35.24 | 3.25 |
| Myalgia (51) | 5 (1.5) | 0 (0.25) | 55.96 | 3.82 |
| Nerve pain (46) | 5 (1.25) | 0 (1) | 46.14 | 3.52 |
| Paraesthesia (36) | 4 (0.5) | 2 (0.88) | 38.90 | 3.92 |
| Poor concentration (49) | 6 (0.5) | 0 (0.75) | 43.66 | 3.67 |
| Tinnitus (25) | 5 (0.5) | 3 (0.5) | 38.57 | 4.72 |
| Fatigue (75) | 7 (0.5) | 5 (1) | 36.03 | 3.69 |
| Nominal aphasia (44) | 6 (0.5) | 0.5 (1) | 34.22 | 3.89 |
| Overall wellbeing (98) | 5 (0.5) | 5 (0.5) | 16.43 | 5.93 |

Symptom severity was rated on a scale from 0 to 10, where 0 indicates no symptoms, 1 indicates mild symptoms, and 10 indicates the most severe symptoms experienced.

^a^Percentage difference (% Diff) represents the relative reduction in median symptom severity following vitamin B12 treatment compared with baseline, calculated using individualised response windows.

^b^Mean response duration represents the average number of days following injection during which symptom severity remained below baseline or within the defined response threshold, reflecting the duration of symptomatic relief.

^c^Values in parentheses following each symptom name (e.g., Dizziness (20)) indicate the total number of injection events contributing to that symptom analysis, not unique participants. Individual participants may therefore contribute multiple injection events.

**Supplementary Table 5.** Pre- and Post-Treatment Symptom and Domain Responses Stratified by Vitamin B_12_ Injection Frequency (3-Day Pre–Post Windows).

| **Outcome** | **Treatment frequency** | **Pre Median (IQR)** | **Post Median (IQR)** | **Δ** | **% Change** | **Count** | **Total injection events** |
| --- | --- | --- | --- | --- | --- | --- | --- |
| **Dizziness** | 2–3 Monthly | 4.71 (3.21) | 3.78 (3.44) | –0.93 | –19.8% | 10 | 20 |
| **Dizziness** | Monthly | 2.21 (3.71) | 0.5 (2.44) | –1.71 | –77.4% | 8 | 13 |
| **Dizziness** | Every 2-3 weeks | 2.5 (0.12) | 0 (0.5) | –2.5 | –100% | 3 | 5 |
| **Dizziness** | Weekly or more often | 3.33 (3.07) | 0 (1.56) | –3.33 | –100% | 5 | 20 |
| **Intestinal problems** | 2–3 Monthly | 3 (3) | 2.44 (3.94) | –0.56 | –18.5% | 17 | 35 |
| **Intestinal problems** | Monthly | 5 (4.33) | 4.67 (6.23) | –0.33 | –6.7% | 9 | 23 |
| **Intestinal problems** | Every 2-3 weeks | 5.87 (1.55) | 3.37 (2.08) | –2.5 | –42.6% | 4 | 12 |
| **Intestinal problems** | Weekly or more often | 5.83 (2.48) | 6.04 (3.25) | 0.21 | 3.6% | 6 | 32 |
| **Irritability** | 2–3 Monthly | 4 (3.58) | 4.17 (3.74) | 0.17 | 4.2% | 15 | 29 |
| **Irritability** | Monthly | 3.83 (1.25) | 1.5 (4.24) | –2.33 | –60.9% | 7 | 14 |
| **Irritability** | Every 2-3 weeks | 3 (1.33) | 0 (0) | –3 | –100% | 2 | 5 |
| **Irritability** | Weekly or more often | 3.75 (0.54) | 2.42 (1.56) | –1.33 | –35.6% | 6 | 26 |
| **Memory loss** | 2–3 Monthly | 4.42 (2.81) | 2.33 (4.17) | –2.08 | –47.2% | 15 | 24 |
| **Memory loss** | Monthly | 4.48 (1.84) | 3.27 (2.68) | –1.21 | –27% | 6 | 18 |
| **Memory loss** | Every 2-3 weeks | 4.33 (2.2) | 3.93 (2) | –0.4 | –9.2% | 3 | 10 |
| **Memory loss** | Weekly or more often | 3.22 (2.55) | 1.83 (1.88) | –1.38 | –43% | 6 | 31 |
| **Myalgia** | 2–3 Monthly | 5 (1.63) | 3.83 (3.44) | –1.17 | –23.3% | 15 | 30 |
| **Myalgia** | Monthly | 4.75 (3.72) | 2.5 (3.03) | –2.25 | –47.4% | 13 | 33 |
| **Myalgia** | Every 2-3 weeks | 4.56 (3.07) | 2.53 (6.01) | –2.03 | –44.5% | 6 | 17 |
| **Myalgia** | Weekly or more often | 7.5 (1.95) | 4.82 (3.88) | –2.68 | –35.7% | 7 | 47 |
| **Nerve pain** | 2–3 Monthly | 5.33 (2) | 4 (2.44) | –1.33 | –25% | 15 | 35 |
| **Nerve pain** | Monthly | 6 (2.49) | 4.11 (3.49) | –1.89 | –31.5% | 7 | 19 |
| **Nerve pain** | Every 2-3 weeks | 4.17 (1.15) | 2.14 (4.36) | –2.03 | –48.7% | 4 | 9 |
| **Nerve pain** | Weekly or more often | 2.98 (3.36) | 2.22 (4.29) | –0.76 | –25.4% | 6 | 28 |
| **Paraesthesia** | 2–3 Monthly | 3.25 (3.33) | 2 (4.67) | –1.25 | –38.5% | 17 | 31 |
| **Paraesthesia** | Monthly | 4.06 (3.52) | 2.64 (3.1) | –1.42 | –34.9% | 8 | 22 |
| **Paraesthesia** | Every 2-3 weeks | 3.67 (2.84) | 3.08 (2.34) | –0.59 | –16.1% | 4 | 13 |
| **Paraesthesia** | Weekly or more often | 4.25 (2.07) | 3.5 (2.77) | –0.75 | –17.6% | 7 | 33 |
| **Poor concentration** | 2–3 Monthly | 4.83 (3.58) | 2.17 (3.79) | –2.67 | –55.2% | 18 | 32 |
| **Poor concentration** | Monthly | 4.58 (2.11) | 3.08 (3.3) | –1.5 | –32.7% | 7 | 20 |
| **Poor concentration** | Every 2-3 weeks | 4 (0.28) | 0.5 (2.2) | –3.5 | –87.5% | 5 | 12 |
| **Poor concentration** | Weekly or more often | 5 (2.46) | 2.62 (3.11) | –2.38 | –47.7% | 8 | 39 |
| **Tinnitus** | 2–3 Monthly | 5.33 (1.17) | 4 (5) | –1.33 | –25% | 13 | 22 |
| **Tinnitus** | Monthly | 5.83 (6.33) | 2.6 (4.22) | –3.24 | –55.5% | 9 | 23 |
| **Tinnitus** | Every 2-3 weeks | 5.55 (1.99) | 3.58 (1.79) | –1.97 | –35.4% | 4 | 10 |
| **Tinnitus** | Weekly or more often | 3.37 (2.66) | 3.2 (1.52) | –0.17 | –5% | 6 | 31 |
| **Fatigue** | 2–3 Monthly | 6.67 (2.35) | 3.25 (5.04) | –3.42 | –51.3% | 28 | 53 |
| **Fatigue** | Monthly | 6.31 (2.54) | 2.78 (5.25) | –3.53 | –55.9% | 14 | 43 |
| **Fatigue** | Every 2-3 weeks | 5.78 (2.36) | 3.28 (3.72) | –2.5 | –43.3% | 6 | 19 |
| **Fatigue** | Weekly or more often | 6.61 (2.33) | 5.61 (2.82) | –1 | –15.1% | 6 | 46 |
| **Nominal aphasia** | 2–3 Monthly | 4.12 (4.08) | 2.5 (3.83) | –1.62 | –39.4% | 12 | 21 |
| **Nominal aphasia** | Monthly | 5.5 (2.15) | 3.44 (1.19) | –2.06 | –37.4% | 7 | 22 |
| **Nominal aphasia** | Every 2-3 weeks | 4.33 (2.13) | 3.5 (1.97) | –0.83 | –19.2% | 3 | 7 |
| **Nominal aphasia** | Weekly or more often | 3.93 (2.22) | 1.15 (2.43) | –2.78 | –70.8% | 6 | 33 |
| **Global Symptom Burden** | 2–3 Monthly | 4.58 (1.48) | 2.79 (2.79) | –1.79 | –39% | 32 | 63 |
| **Global Symptom Burden** | Monthly | 4.74 (2.19) | 2.53 (1.74) | –2.2 | –46.5% | 16 | 51 |
| **Global Symptom Burden** | Every 2–3 weeks | 4.48 (1.53) | 2.99 (2.87) | –1.49 | –33.3% | 6 | 20 |
| **Global Symptom Burden** | Weekly or more often | 4.94 (2.09) | 3.1 (1.41) | –1.84 | –37.3% | 8 | 53 |
| **Overall wellbeing** | 2–3 Monthly | 5 (2.92) | 4.17 (4.26) | -0.83 | -16.7% | 31 | 63 |
| **Overall wellbeing** | Monthly | 5.39 (2.08) | 3.06 (2.59) | -1.55 | -28.9% | 16 | 53 |
| **Overall wellbeing** | Every 2-3 weeks | 5.81 (1.39) | 3.92 (1.17) | -0.27 | -4.8% | 6 | 20 |
| **Overall wellbeing** | Weekly or more often | 5.07 (3) | 3.25 (1.94) | -1.68 | -33.1% | 7 | 51 |
| **Cognitive mean** | 2–3 Monthly | 2.42 (2.92) | 1.11 (2.1) | –1.31 | –54% | 22 | 40 |
| **Cognitive mean** | Monthly | 3.17 (3.63) | 2.11 (2.16) | –1.06 | –33.3% | 9 | 26 |
| **Cognitive mean** | Every 2-3 weeks | 1.33 (1.83) | 0.89 (2.61) | –0.44 | –33.3% | 5 | 13 |
| **Cognitive mean** | Weekly or more often | 2.51 (1.33) | 2.05 (1.95) | –0.45 | –18% | 8 | 43 |
| **Neuro-sensory domain** | 2–3 Monthly | 1.62 (2.12) | 1.25 (2.33) | –0.38 | –23.1% | 26 | 50 |
| **Neuro-sensory domain** | Monthly | 1.96 (2.17) | 1.22 (2.45) | –0.74 | –37.6% | 15 | 43 |
| **Neuro-sensory domain** | Every 2-3 weeks | 2.29 (1.44) | 1.85 (1.9) | –0.45 | –19.5% | 6 | 19 |
| **Neuro-sensory domain** | Weekly or more often | 1.74 (3.3) | 1.44 (3.15) | –0.3 | –17.4% | 8 | 46 |
| **Pain-fatigue-affective domain** | 2–3 Monthly | 3.17 (3.11) | 2.31 (2.36) | –0.85 | –26.9% | 31 | 57 |
| **Pain-fatigue-affective domain** | Monthly | 2.56 (2.45) | 1.37 (2.55) | –1.19 | –46.4% | 15 | 48 |
| **Pain-fatigue-affective domain** | Every 2-3 weeks | 3.76 (1.7) | 2.05 (3.2) | –1.71 | –45.6% | 6 | 20 |
| **Pain-fatigue-affective domain** | Weekly or more often | 4.42 (1.45) | 3.98 (0.86) | –0.44 | –9.9% | 7 | 48 |

Note. Symptom severity was rated on a 0–10 scale (0 = none; 10 = most severe). Values are medians (IQR) of participant-level averages, where injection-level pre/post means were first averaged within each participant to avoid over-representation of frequent injectors. Pre-treatment values represent the mean symptom severity across the 3 days immediately prior to injection; post-treatment values represent the mean symptom severity across the 3 days immediately following injection. % change was calculated as ((Median_post - Median_pre) / Median_pre) × 100. Count indicates the number of participants contributing paired values in that frequency group; Total injection events indicates the number of eligible isolated injection events contributing to that estimate. GlobalSymptomBurden reflects the mean severity across symptoms experienced on a given day (excluding overall wellbeing). Domain scores (cognitive_mean, neuro_mean, painfatigue_mean) represent daily mean severity across symptoms within each domain.

**Supplementary Table 6.** Symptom-specific treatment response phenotypes with pre- and post-treatment median severity and absolute change following vitamin B_12_ injection (n = 62).

| **Symptom** | **Response Category** | **Count** | **Percentage** | **Pre Median (IQR)** | **Post Median (IQR)** | **Absolute change** |
| --- | --- | --- | --- | --- | --- | --- |
| **Dizziness** | Substantial Improvement | 16 | 47.1 | 2.58 (1.88) | 0.00 (1.00) | 2.58 |
| **Dizziness** | Moderate Improvement | 2 | 5.9 | 4.86 (2.36) | 3.19 (1.53) | 1.67 |
| **Dizziness** | Minimal Improvement | 2 | 5.9 | 4.28 (0.78) | 3.35 (0.52) | 0.93 |
| **Dizziness** | Non-Responder | 3 | 8.8 | 8.17 (2.33) | 7.67 (2.33) | 0.5 |
| **Dizziness** | Worsening | 3 | 8.8 | 5.67 (1.46) | 6.25 (2.12) | -0.58 |
| **Dizziness** | Symptom Only Present After Treatment | 8 | 23.5 | 0.00 (0.00) | 2.17 (2.08) | -2.17 |
| **Intestinal problems** | Substantial Improvement | 13 | 31.7 | 2.67 (2.78) | 0.00 (0.00) | 2.67 |
| **Intestinal problems** | Moderate Improvement | 5 | 12.2 | 6.06 (0.40) | 3.94 (0.27) | 2.11 |
| **Intestinal problems** | Minimal Improvement | 7 | 17.1 | 6.92 (2.70) | 5.33 (2.53) | 1.58 |
| **Intestinal problems** | Non-Responder | 5 | 12.2 | 6.30 (2.25) | 6.23 (1.75) | 0.07 |
| **Intestinal problems** | Worsening | 6 | 14.6 | 4.04 (2.98) | 5.42 (3.19) | -1.38 |
| **Intestinal problems** | Symptom Only Present After Treatment | 5 | 12.2 | 0.00 (0.00) | 2.42 (1.67) | -2.42 |
| **Irritability** | Substantial Improvement | 12 | 35.3 | 3.00 (1.31) | 0.20 (0.71) | 2.8 |
| **Irritability** | Moderate Improvement | 4 | 11.8 | 4.00 (0.75) | 2.42 (0.64) | 1.58 |
| **Irritability** | Minimal Improvement | 7 | 20.6 | 7.00 (2.12) | 5.33 (1.50) | 1.67 |
| **Irritability** | Non-Responder | 5 | 14.7 | 3.50 (3.83) | 3.50 (4.00) | 0 |
| **Irritability** | Worsening | 2 | 5.9 | 4.33 (0.67) | 5.67 (1.33) | -1.33 |
| **Irritability** | Symptom Only Present After Treatment | 4 | 11.8 | 0.00 (0.00) | 4.25 (2.31) | -4.25 |
| **Memory loss** | Substantial Improvement | 14 | 42.4 | 4.17 (2.94) | 0.44 (1.79) | 3.72 |
| **Memory loss** | Moderate Improvement | 3 | 9.1 | 3.43 (1.58) | 1.83 (1.05) | 1.6 |
| **Memory loss** | Minimal Improvement | 2 | 6.1 | 4.33 (1.67) | 3.50 (1.17) | 0.83 |
| **Memory loss** | Non-Responder | 9 | 27.3 | 4.46 (1.00) | 4.56 (1.50) | -0.1 |
| **Memory loss** | Worsening | 2 | 6.1 | 4.42 (1.08) | 6.00 (0.50) | -1.58 |
| **Memory loss** | Symptom Only Present After Treatment | 3 | 9.1 | 0.00 (0.00) | 1.00 (0.08) | -1 |
| **Myalgia** | Substantial Improvement | 15 | 35.7 | 4.67 (3.67) | 0.00 (1.47) | 4.67 |
| **Myalgia** | Moderate Improvement | 3 | 7.1 | 6.33 (1.94) | 4.33 (1.44) | 2 |
| **Myalgia** | Minimal Improvement | 15 | 35.7 | 5.22 (2.58) | 4.25 (2.59) | 0.97 |
| **Myalgia** | Non-Responder | 4 | 9.5 | 5.79 (0.39) | 5.38 (0.44) | 0.42 |
| **Myalgia** | Worsening | 4 | 9.5 | 5.31 (1.60) | 6.17 (1.54) | -0.86 |
| **Myalgia** | Symptom Only Present After Treatment | 1 | 2.4 | 0.00 (0.00) | 2.00 (0.00) | -2 |
| **Nerve pain** | Substantial Improvement | 8 | 23.5 | 3.75 (2.38) | 0.00 (0.12) | 3.75 |
| **Nerve pain** | Moderate Improvement | 6 | 17.6 | 5.75 (1.53) | 3.55 (1.14) | 2.2 |
| **Nerve pain** | Minimal Improvement | 6 | 17.6 | 5.42 (0.70) | 4.33 (0.93) | 1.08 |
| **Nerve pain** | Non-Responder | 9 | 26.5 | 6.00 (2.67) | 5.75 (2.06) | 0.25 |
| **Nerve pain** | Worsening | 3 | 8.8 | 2.46 (2.00) | 3.17 (2.17) | -0.71 |
| **Nerve pain** | Symptom Only Present After Treatment | 2 | 5.9 | 0.00 (0.00) | 5.83 (0.17) | -5.83 |
| **Paraesthesia** | Substantial Improvement | 12 | 31.6 | 2.17 (2.19) | 0.00 (0.25) | 2.17 |
| **Paraesthesia** | Moderate Improvement | 7 | 18.4 | 3.25 (3.08) | 2.00 (2.08) | 1.25 |
| **Paraesthesia** | Minimal Improvement | 7 | 18.4 | 4.50 (2.36) | 3.33 (1.59) | 1.17 |
| **Paraesthesia** | Non-Responder | 8 | 21.1 | 5.75 (3.10) | 5.46 (3.03) | 0.29 |
| **Paraesthesia** | Worsening | 2 | 5.3 | 1.42 (0.08) | 1.67 (-0.00) | -0.25 |
| **Paraesthesia** | Symptom Only Present After Treatment | 2 | 5.3 | 0.00 (0.00) | 1.17 (0.50) | -1.17 |
| **Poor concentration** | Substantial Improvement | 18 | 43.9 | 4.29 (3.26) | 0.00 (1.04) | 4.29 |
| **Poor concentration** | Moderate Improvement | 8 | 19.5 | 4.52 (2.21) | 2.83 (1.18) | 1.69 |
| **Poor concentration** | Minimal Improvement | 3 | 7.3 | 4.67 (0.88) | 4.00 (0.46) | 0.67 |
| **Poor concentration** | Non-Responder | 7 | 17.1 | 7.11 (3.31) | 6.78 (3.84) | 0.33 |
| **Poor concentration** | Worsening | 2 | 4.9 | 3.00 (2.33) | 3.62 (2.62) | -0.62 |
| **Poor concentration** | Symptom Only Present After Treatment | 3 | 7.3 | 0.00 (0.00) | 1.33 (2.33) | -1.33 |
| **Tinnitus** | Substantial Improvement | 12 | 37.5 | 1.58 (5.29) | 0.00 (0.92) | 1.58 |
| **Tinnitus** | Moderate Improvement | 5 | 15.6 | 6.00 (0.39) | 3.67 (0.50) | 2.33 |
| **Tinnitus** | Minimal Improvement | 4 | 12.5 | 4.97 (0.82) | 4.00 (1.08) | 0.97 |
| **Tinnitus** | Non-Responder | 10 | 31.2 | 5.79 (2.79) | 5.33 (2.91) | 0.46 |
| **Tinnitus** | Worsening | 1 | 3.1 | 1.33 (0.00) | 3.00 (0.00) | -1.67 |
| **Fatigue** | Substantial Improvement | 20 | 36.4 | 5.50 (2.92) | 1.17 (2.06) | 4.33 |
| **Fatigue** | Moderate Improvement | 4 | 7.3 | 6.86 (1.25) | 4.81 (1.12) | 2.06 |
| **Fatigue** | Minimal Improvement | 9 | 16.4 | 5.67 (2.67) | 4.22 (1.56) | 1.44 |
| **Fatigue** | Non-Responder | 18 | 32.7 | 6.82 (1.40) | 6.65 (0.97) | 0.18 |
| **Fatigue** | Worsening | 3 | 5.5 | 7.00 (1.56) | 9.33 (1.33) | -2.33 |
| **Fatigue** | Symptom Only Present After Treatment | 1 | 1.8 | 0.00 (0.00) | 2.00 (0.00) | -2 |
| **Nominal aphasia** | Substantial Improvement | 13 | 44.8 | 3.50 (3.00) | 0.67 (1.33) | 2.83 |
| **Nominal aphasia** | Moderate Improvement | 4 | 13.8 | 6.38 (2.19) | 4.13 (1.31) | 2.25 |
| **Nominal aphasia** | Minimal Improvement | 4 | 13.8 | 5.17 (0.67) | 4.00 (0.25) | 1.17 |
| **Nominal aphasia** | Non-Responder | 6 | 20.7 | 4.92 (3.10) | 4.75 (2.90) | 0.17 |
| **Nominal aphasia** | Worsening | 1 | 3.4 | 1.33 (0.00) | 3.11 (0.00) | -1.78 |
| **Nominal aphasia** | Symptom Only Present After Treatment | 1 | 3.4 | 0.00 (0.00) | 1.42 (0.00) | -1.42 |
| **Global Symptom Burden** | Substantial Improvement | 18 | 29 | 3.64 (2.01) | 1.42 (1.03) | 2.22 |
| **Global Symptom Burden** | Moderate Improvement | 13 | 21 | 4.85 (2.23) | 2.75 (1.14) | 2.1 |
| **Global Symptom Burden** | Minimal Improvement | 13 | 21 | 4.71 (1.74) | 3.94 (1.52) | 0.77 |
| **Global Symptom Burden** | Non-Responder | 11 | 17.7 | 4.66 (1.24) | 4.33 (1.37) | 0.33 |
| **Global Symptom Burden** | Worsening | 7 | 11.3 | 2.04 (1.34) | 2.46 (1.89) | -0.41 |
| **Cognitive domain** | Substantial Improvement | 23 | 47.9 | 2.17 (2.73) | 0.56 (1.33) | 1.61 |
| **Cognitive domain** | Moderate Improvement | 5 | 10.4 | 1.33 (1.33) | 0.89 (0.50) | 0.44 |
| **Cognitive domain** | Minimal Improvement | 3 | 6.2 | 3.75 (1.42) | 2.83 (1.01) | 0.92 |
| **Cognitive domain** | Non-Responder | 8 | 16.7 | 4.56 (1.82) | 4.22 (2.03) | 0.35 |
| **Cognitive domain** | Worsening | 5 | 10.4 | 1.83 (0.79) | 2.17 (0.76) | -0.33 |
| **Cognitive domain** | Symptom Only Present After Treatment | 4 | 8.3 | 0.00 (0.00) | 0.49 (0.62) | -0.49 |
| **Neuro-sensory domain** | Substantial Improvement | 12 | 20.7 | 1.04 (0.51) | 0.00 (0.17) | 1.04 |
| **Neuro-sensory domain** | Moderate Improvement | 14 | 24.1 | 1.73 (1.54) | 1.00 (0.88) | 0.73 |
| **Neuro-sensory domain** | Minimal Improvement | 11 | 19 | 2.00 (2.78) | 1.50 (2.33) | 0.5 |
| **Neuro-sensory domain** | Non-Responder | 16 | 27.6 | 2.83 (2.10) | 2.80 (2.17) | 0.03 |
| **Neuro-sensory domain** | Worsening | 2 | 3.4 | 2.00 (0.25) | 2.54 (0.04) | -0.54 |
| **Neuro-sensory domain** | Symptom Only Present After Treatment | 3 | 5.2 | 0.00 (0.00) | 0.58 (0.23) | -0.58 |
| **Pain-fatigue-affective domain** | Substantial Improvement | 14 | 23.3 | 2.31 (1.86) | 0.41 (0.68) | 1.91 |
| **Pain-fatigue-affective domain** | Moderate Improvement | 12 | 20 | 3.92 (2.92) | 2.49 (1.67) | 1.43 |
| **Pain-fatigue-affective domain** | Minimal Improvement | 14 | 23.3 | 4.00 (2.58) | 3.23 (2.02) | 0.77 |
| **Pain-fatigue-affective domain** | Non-Responder | 15 | 25 | 4.11 (2.69) | 4.17 (2.75) | -0.06 |
| **Pain-fatigue-affective domain** | Worsening | 4 | 6.7 | 2.61 (1.53) | 3.68 (1.65) | -1.07 |
| **Pain-fatigue-affective domain** | Symptom Only Present After Treatment | 1 | 1.7 | 0.00 (0.00) | 0.50 (0.00) | -0.5 |

**Note:** Symptom severity was recorded daily on an 11-point scale (0–10; higher scores indicate greater severity) before and after vitamin B12 injections in people with pernicious anaemia. Participants were classified into symptom-specific treatment response categories based on changes between the pre- and post-treatment windows, using a hybrid framework that combined absolute point change and percentage change thresholds (substantial, moderate, minimal improvement, non-response, worsening). Pre- and post-treatment severity is summarised as median (interquartile range, IQR). Absolute change represents the difference between pre- and post-treatment medians (Pre − Post), with positive values indicating improvement. Counts indicate the number of participants per response category, and percentages are calculated within each symptom. The category “Symptom Only Present After Treatment” denotes symptoms absent pre-treatment but reported post-treatment and is included for completeness rather than to indicate treatment response.
